# Functional Heterogeneity in Non-Suicidal Self-Injury across Psychiatric Disorders: Neural and Psychosocial Correlates

**DOI:** 10.1101/2025.01.23.25320985

**Authors:** Mingzhu Li, Yang Xiao, Yuqi Ge, Huiru Yan, Xueni Li, Weihua Yue, Hao Yan

## Abstract

Non-suicidal self-injury (NSSI) is a common behavior among adolescents, particularly within psychiatric populations. While neurobiological and psychosocial risk factors have been extensively studied, the mechanisms underlying NSSI’s heterogeneity remain unclear. This study investigated 304 hospitalized adolescents/young adults (16–25 years) with NSSI and comorbid psychiatric diagnoses (major depressive disorder [MDD], bipolar disorder [BD], eating disorders [ED]) using psychological assessments and resting-state fMRI data from 163 participants. Orthogonal projection non-negative matrix factorization of Ottawa Self-Injury Inventory responses identified two core motivational dimensions: self-related (emotional regulation) and social-related (interpersonal influence). The self-related factor correlated with amygdala-centered cortico-limbic emotional regulation networks and predominated in affective disorders (MDD/BD), while the social-related factor linked to frontoparietal cognitive control and frontotemporal social cognition networks, particularly in ED. Fuzzy C-means clustering revealed three NSSI functional subtypes, independent of diagnostic categories : self-subtype primarily driven by self-related functions, social-subtype influenced by both self- and social-related functions with greater exposure to psychosocial risks, and non-specific subtype with undifferentiated motivations. No subtype was exclusively driven by social-related functions. The “self-social” dual-dimensional neural model demonstrated distinct subtype-specific profiles in functional connectivity, psychosocial risk exposure, and clinical features. Self-related mechanisms primarily engaged emotional regulation circuits, whereas social-related mechanisms emphasize the role of psychosocial risk factors and cognitive-emotional circuits. These findings provide neural evidence for the functional heterogeneity of NSSI and highlight the need for personalized interventions. Treatments targeting emotion regulation may benefit all subtypes, individuals with prominent social-related motivations may additionally require interventions aimed at improving interpersonal functioning.

## Introduction

Non-suicidal self-injury (NSSI), defined as the deliberate self-inflicted damage to body tissues without suicidal intent (e.g., cutting, burning, or hitting) ^1^, is a prevalent behavior among adolescents and young adults, particularly in psychiatric populations ^2^. Lifetime prevalent rates of NSSI range from 13.4–17.2% in community samples ^3^ to as high as 67% in psychiatric inpatient settings ^4^. NSSI frequently co-occurs with major depressive disorders (MDD), bipolar disorders (BD), and eating disorders (ED) ^5^, underscoring its transdiagnostic nature. Although not intended to be suicidal, NSSI is associated with severe adverse outcomes, including increased suicide risk ^6^ and impaired social functioning ^7^, underscoring its significant public health impact.

NSSI is a complex behavior shaped by the interplay of social, psychological, and biological factors ^8^. Psychosocial risk variables, such as childhood abuse, adverse life events, limited family and social support, depression, anxiety and deficits in emotion regulation are well-documented contributors to NSSI ^9–11^. These variables can be broadly categorized into interpersonal and intrapersonal domains, both of which play critical roles in the onset and maintenance of NSSI ^12^. Although advances in psychosocial research have successfully informed emotion regulation-focused psychological interventions ^13^, the lack of mechanistic elucidation at the biological level continues to hinder treatment innovation. To date, no targeted somatic treatments, such as psychopharmacology or neuromodulation, are available for NSSI ^14^, highlighting the urgent need to elucidate its neurobiological mechanisms ^15^.

From a neurodevelopmental perspective, distal risk factors (e.g., childhood trauma) during critical periods of neural maturation can influence proximal biological alterations, including structural and functional changes in key brain regions ^16^. Neuroimaging studies investigating the neural correlates of NSSI have identified abnormalities in the frontal-limbic system, encompassing the amygdala, anterior cingulate cortex (ACC), medial prefrontal cortex (mPFC), dorsolateral prefrontal cortex (dlPFC), hippocampus, and insula ^17,18^. These regions are integral to emotion expression and regulation ^19^, self-referential processing/interpersonal interactions ^20^, and reward processing ^21^. For instance, hyperactivity in the amygdala and insula has been linked to heightened emotional reactivity and salience detection ^19^, while dysregulation in the mPFC and ACC is associated with impaired self-regulation and social cognition ^20^. These findings suggest that NSSI may arise from dysregulated bottom-up and top-down neural circuits involved in emotion and cognitive control ^17^.

NSSI is a highly heterogeneous behavior, varying in frequency, methods, severity, and underlying motivations ^22^. The functional model of NSSI, which distinguishes between intrapersonal (e.g., emotion regulation) and interpersonal (e.g., emotion expression and social influence) functions, provides a valuable framework for understanding this heterogeneity ^23,24^. Studies indicate that 66–81% of individuals engage in NSSI primarily for intrapersonal functions, while 33–56% report interpersonal motivations ^25^. These functional dimensions are associated with differing clinical outcomes, with intrapersonal functions linked to higher suicide risk ^26,27^. Moreover, inter- and intrapersonal functions appear to be differentially associated with abovementioned psychosocial factors and neurobiological mechanisms, respectively.

In this study, we aim to bridge the gap between self-injurious behaviors and their underlying neurobiological and psychosocial correlates. First, we identify conceptually meaningful and compact factor structures for NSSI functions in hospitalized adolescents with psychiatric disorders and derived functional subtypes. Next, we examined the neural correlates of different factors of NSSI functions. Finally, we explored the factor related neural characteristics and psychosocial risk variables associated with the different functional subtypes. By integrating functional, neural, and psychosocial dimensions, this study seeks to advance our understanding of NSSI mechanisms and inform the development of targeted psychological and neuromodulation interventions.

## Methods

### 2.1 Participants

The present study included inpatients at Peking University Sixth Hospital (Beijing, Chian) from July 2021 to September 2022. A total of 304 patients were enrolled and completed the full set of questionnaires. Among them, 163 patients voluntarily underwent magnetic resonance imaging (MRI) data acquisition. This study was approved by the Ethics Committee of Peking University Sixth Hospital.

Inclusion criteria were: (1) aged 16-25 years, of any gender; (2) inpatient diagnosis meeting the International Classification of Diseases, 10th Revision (ICD-10) criteria for depressive episode or recurrent depressive disorder, bipolar disorder, and eating disorders, as assessed by two attending or senior psychiatrists; (3) at least one episode of NSSI; (4) provision of written informed consent by the patient or their guardian.

Exclusion criteria were: (1) individuals exhibiting only suicidal behavior without NSSI; (2) those with mental developmental delay; (3) individuals with severe, unstable physical illnesses; (4) history of epilepsy or febrile seizures; (5) pregnant or lactating women, or those planning pregnancy.

Among the participants, 167 were diagnosed with major depressive disorders, 89 with bipolar disorders, and 48 with eating disorders. This study included patients with varying severities of NSSI behaviors: 178 participants had repeated NSSI behaviors (NSSI behaviors occurred for at least 5 days in the past year, meeting the DSM-5 diagnostic criteria for NSSI disorder), while 126 patients had occasional NSSI behaviors (NSSI behaviors occurred on at least one day but fewer than 5 days). Given that the focus of this study is on the functions of NSSI behaviors, the functional classifications were applied to both frequent and occasional NSSI behaviors to maintain consistency across varying severities.

### 2.2 Measurements and Assessments

Basic demographic characteristics, medical history, comorbidities, and clinical information regarding current psychiatric disorders were collected using a self-designed Case Report Form (CRF). Suicidal ideation and suicidal behavior were also recorded in the CRF based on documentation in the patients’ hospitalization records regarding the presence of suicidal ideation and suicidal behavior prior to the current admission.

The Ottawa Self-Injury Inventory (OSI) was used to assess the frequency and functions of NSSI behaviors among participants. This scale has demonstrated good reliability and validity in Chinese adolescent samples ^28^. The study specifically focused on the NSSI functions assessed by the 29 items of the OSI function subscale (OSI-F) ^28^, which captures various motivations and intentions underlying NSSI behaviors. Responses were rated from 0 (never) to 4 (always), with higher scores indicating stronger functional tendencies.

The Self-Rating Anxiety Scale (SAS) ^29^ and the Self-Rating Depression Scale (SDS) ^29^ were used to evaluate anxiety and depressive symptoms, respectively. Additionally, several other scales were utilized to assess individual characteristics and environmental risk factors, including the NEO Five-Factor Inventory (NEOFFI) ^30^, the revised Chinese Internet Addiction Scale (CIAS-R) ^31^, the Adolescent Self-Rating Life Events Check List (ASLEC) ^29^, the Childhood Trauma Questionnaire (CTQ) ^32^, the Family Assessment Device (FAD) ^29^, and the Social Support Rate Scale (SSRS) ^29^. Suicidal ideation and suicide attempt within the past year were also recorded.

### 2.3 Factor Analysis of NSSI Functions

Orthonormal projective non-negative factorization (OPNMF) ^33,34^ was employed to identify latent factors underlying the NSSI functional items from the OSI-F. OPNMF is a variant of NMF, which is widely used in recent biomedical studies^35^. This method introduces two constraints: a projective constraint, which ensures stable and deterministic item-to-factor assignments, and an orthonormality constraint, which promotes compact and non-redundant factor definitions. Here, the OPNMF achieved factorization for given data (OSI-F) by yielding a probabilistic parcellation to assign latent factor loadings for each item of NSSI functions (a dictionary matrix containing factors), and these loadings quantified the extent of which each item belongs to a factor. The advantages of OPNMF enabled us to acquire a sparse, stable and interpretable structure of latent factor of NSSI functions.

To determine the optimal number of factors, we employed a set of sophisticated evaluation strategies based on cross-validation with 10,000 split-half analyses. Specifically, the entire sample used in the current study was divided into two halves, and the OPNMF was conducted separately on each half to derive the dictionary matrices. We subsequently calculated the congruency between item-to-factor assignments to examine the stability of the factor solutions at various evaluation strategies (describe in the Supplement), including the adjusted Rand index (RI), variation of information (VI), and concordance index (CI) between the dictionaries ^35^. Additionally, we also assessed the generalizability of the factor model by using the reconstruction error (RE) calculating the absolute differences between the projection of the dictionary from one split-sample data on the other split-sample ^35^. The internal consistency of the optimal OPNMF model was assessed using the Pearson’s correlations with an additional bootstrapping validation procedure in which we conducted 10,000 times resampling and re-calculate the correlation coefficient. The between-items relationships under optimal factor structure were also calculated using the Pearson’s correlation.

### 2.4 Clustering of NSSI Functional Subtypes

Factor loadings from the optimal OPNMF model were used to cluster patients into different NSSI functional subtypes. Considering the extent of differentiation of vulnerability factors related to disorder or symptoms exhibited a certain of heterogeneity in different patients, we adopted a soft clustering approach (Fuzzy C-means) to allow probabilistic assignment to multiple subtypes. This analytic combination has been infrequently explored in NSSI populations.

Fuzzy C-means clustering is a widely used soft clustering algorithm that estimates the membership likelihood of each individual to each cluster, rather than forcing exclusive assignment. This method allows us to identify those patients with an explicit inclination to specific NSSI functions and ambiguous subgroups that have not yet been differentiated into specific categories, based on a cutoff threshold.

In this study, the fuzzy C-means clustering ^36^ was implemented to NSSI factor loadings of all patients to encode the membership likelihood of each patient in any given subtyping clusters, after adjusting for age, gender, years of education, suicidal ideation, and suicide attempts. The fuzzy silhouette index (SI), Xie and Beni index (XB), and partition entropy (PE) were calculated at range of 2 to 6 as internal validity indices to determine the optimal number of clusters (details in Supplement). Higher SI and lower XB and PE indicated better clustering quality. To choose the cutoff to define the optimal membership discrimination, the elbow method was used at range of 70% to 90% by identifying the elbow point, where the rate of decrease in explained variance slows significantly. The elbow point represented a balance between maximizing cluster compactness and minimizing the number of clusters.

### 2.5 Resting-State Functional Connectivity (rsFC) Network Construction

Resting-state functional MRI data were acquired from 163 participants using the Siemens Prisma 3.0 Tesla MRI scanner. Data preprocessing was conducted using the DPABI (Data Processing Assistant for Resting State fMRI Advanced Edition) ^37^ software based on the MATLAB (The MathWorks, Natick, MA, USA). Standard processing procedures were followed. The scanning parameters and detailed information for each preprocessing step are available in the Supplement.

This study quantified resting-state functional connectivity (rsFC) based on the spontaneous fluctuations of resting-state bold oxygenation level dependent (BOLD) signals, generating a whole-brain rsFC matrix for each participant. We referred to a previous brain parcellation scheme comprising a total of 210 cortical regions ^38^ and 14 subcortical regions ^39^. The average time series signals of each brain region were extracted, and the Pearson’s correlation coefficients were computed between the time series signals of each pair of brain regions to determine functional connectivity strength. The Fischer’s *r-to-z* transformation was utilized to convert the correlation coefficients into normalized *z-*scores. A 224 × 224 functional connectivity matrix was constructed for each participant.

### 2.6 Modeling Neural Correlates of NSSI Functions Using rsFC

To better understand the neural correlates of NSSI functions, we trained a machine-learning algorithm based on regularized canonical correlation (RCC) with a hyper-parameterized feature selection space following a previous study ^40^ (details in Supplement).

Briefly, RCC based on L2 penalty (λ) was used to predict each factor loadings of NSSI functions for all patients. Out-of-sample predictions were obtained using a nested cross-validation scheme using a 90% random selection strategy (10-fold), repeated 100 times. During the training of the model in each fold, we ran a random hyperparameter search consisting of 10 iterations in a 10-fold cross-validation using the grid search and chose the best model for prediction. The optimized number of rsFC features was defined as the top-ranked features based on a bootstrapping scheme with 95% subsampling and 100 replications. The hyperparameter space was defined as follows: *N*_features_ ranged from 100 to 400 top-ranked rsFC features, and λ ranged from 1 to 10. We evaluated the performance of each model on the entire predicted set using canonical correlation coefficients. Statistical significance of the coefficients was evaluated using a non-parametric permutation test with 10,000 iterations.

The correlations between the FC scores and each functional factor were using the Pearson correlation analysis. The significant threshold for all statistical analysis was set as *p* < 0.001 adjusted by multiple comparison correction using false discovery rate (FDR) method.

### 2.7 Statistical Analyses

Pearson’s correlation analysis was used to examine the associations between factors loadings and SAS/SDS scores. One-way analysis of variance (ANOVA) with a post-hoc Turkey test was employed to analyze differences in scale scores (e.g., CTQ, ALSE) across different subtypes or diagnostic groups (MDD, BD and ED). For ordinal and categorical variables, such as NSSI frequency and gender, chi-square tests were conducted to assess group differences. The statistical analyses above were conducted using SPSS, version 27 (IBM, Armonk, NY). OPNMF and fuzzy C-means clustering were performed using MATLAB (version 2012a, MathWorks, Natick, MA, USA). All analyses controlled for age, gender, years of education, suicidal ideation, and suicide attempts. A *p* < 0.05 was considered the threshold for statistical significance, multiple comparisons were corrected by FDR method.

## Results

### 3.1 Demographic and Clinical Characteristics

A total of 304 participants were included and completed the full assessment, with 163 of them also undergoing MRI for subsequent analysis of neural correlates. Statistical comparisons indicated that the MRI subset did not significantly differ from the overall sample in terms of age, gender distribution, level of depression and anxiety, personality traits, and various assessments for the environmental risk factors (Table 1). Despite a high proportion of participants in both groups reporting suicidal ideation and attempts, no significant differences were observed (Table 1). Regarding the severity of NSSI, no significant differences were found between the two samples in terms of NSSI frequency, age at first episode, or functional scores (Table 1 and Table S1).

**Figure 1.**
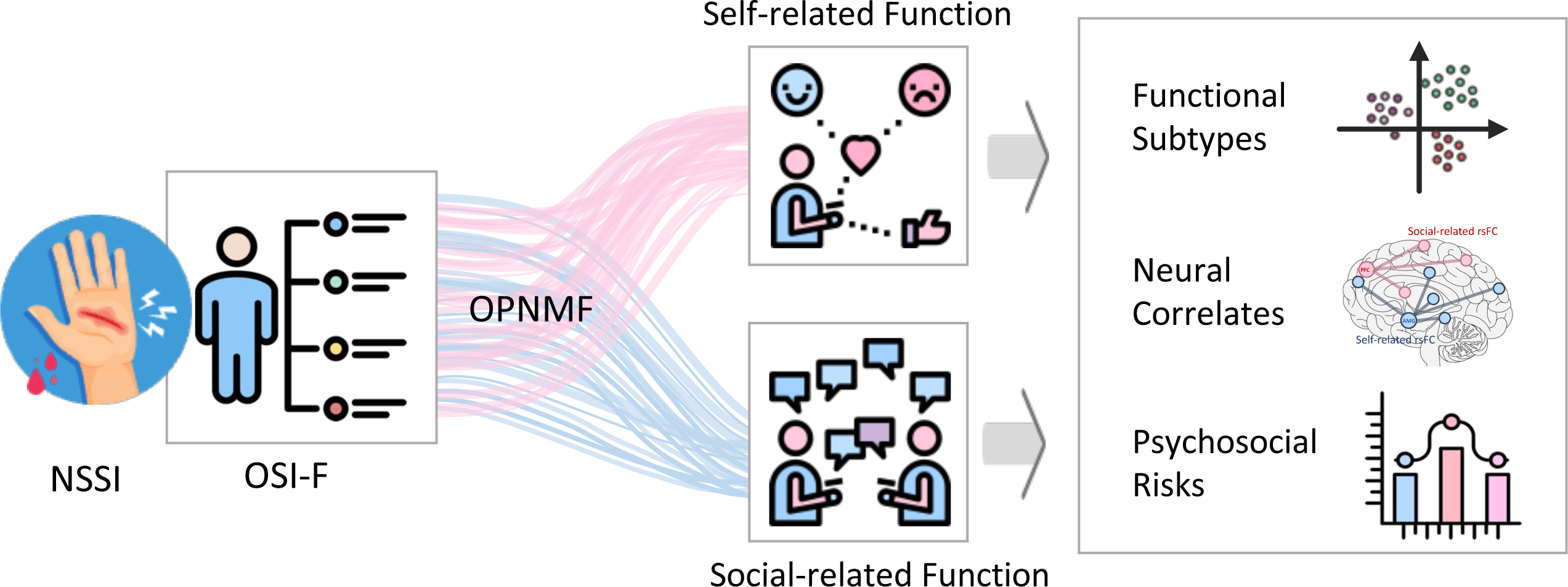
Neurobiological vulnerabilities and psychosocial risks in psychiatric disorders with NSSI. A sample of 304 inpatients diagnosed with major depressive disorder (MDD: N = 167), bipolar disorder (BD: N = 89), and eating disorders (ED: N = 48) reported the functions underlying their NSSI behavior. The self-reported functions were analyzed using orthogonal projection non-negative matrix factorization (OPNMF), which identified two primary functional factors: social-related and self-related functions. Functional connectivity (FC) features and corresponding subtypes were determined based on these factors. Subsequent analyses examined differences in FC characteristics and associated risk factors across the subtypes, offering valuable insights into the neurobiological vulnerabilities and psychosocial risks linked to NSSI behavior. MDD, major depressive disorder; BD, bipolar disorder; ED, eating disorders; OPNMF, orthogonal projection non-negative matrix factorization; OSI-F, functional items in the Ottawa Self-Injury Inventory.

**Table 1.**
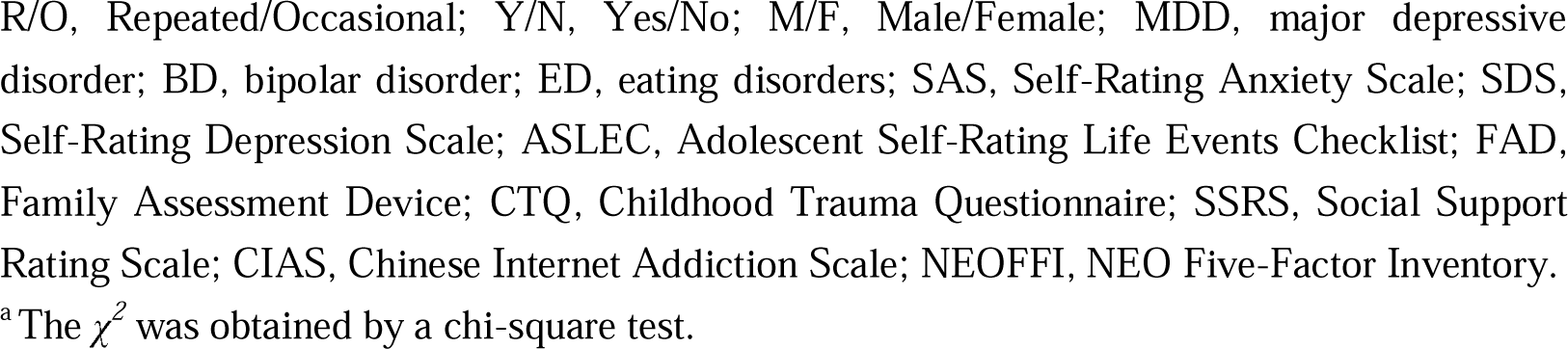
Demographic and clinical characteristics.

### 3.2 Two Factors of NSSI Functions

A robust two-factor model was obtained using OPNMF based on 304 patients (Figure 2A). The first factor comprised 14 NSSI functional items in OSI-F, it mainly related to self-perception and emotional regulation and was named as “self-related factor”. The second factor comprised 15 items mainly related to interpersonal functioning and was named as “social-related factor”. Model evaluation metrics indicated that a two-factor solution provided the highest stability (with the highest adjusted RI and CI values, and the lowest VI value, Figure S1A) and the strongest generalizability (with the lowest out-of-sample increase in RE value, Figure S1A). These results were validated through 1,000 times split-half cross-validation.

**Figure 2.**
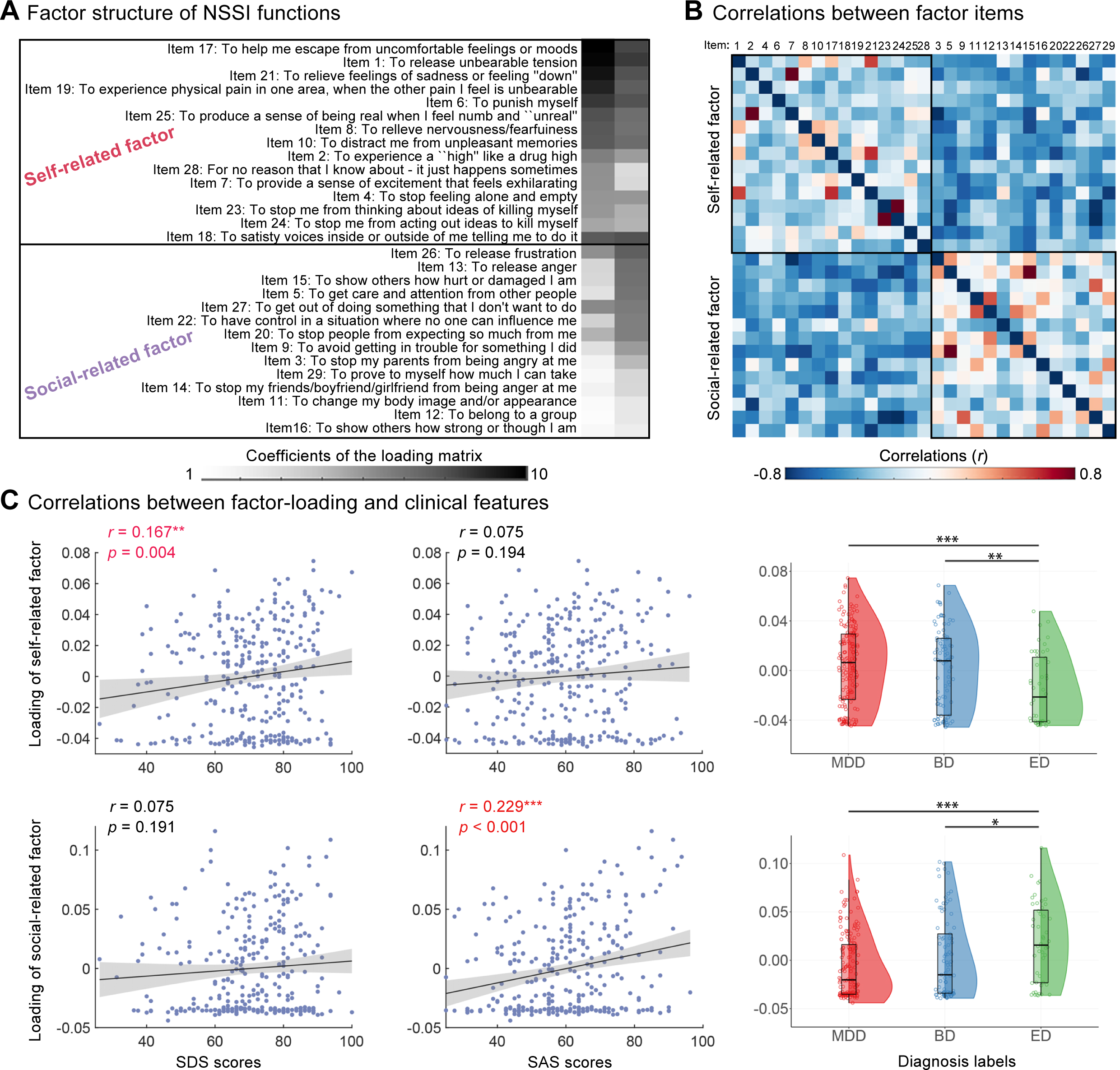
Self–Social dimensional model of NSSI functions. **(A)** The most stable and generalizable two-factor structure of NSSI functions identified by OPNMF (N = 304), consisting of a self-related factor (14 items) and a social-related factor (15 items) derived from the 29 OSI-F items. **(B)** A heatmap illustrating the correlations between and within the two derived factors. The strength of correlations is represented by color: red indicates positive correlations, blue indicates negative correlations, with darker shades denoting stronger correlations. **(C)** Scatter plots display Pearson’s correlations between levels of anxiety and depression and the two factor loadings, adjusted for age, sex, education, SI, and SA. **(D)** One-way ANOVA results comparing the two factor loadings across the three diagnostic groups, controlling for covariates. For each box plot, the box illustrates the Standard Error of the Mean (SEM, centered on the mean), whiskers denote the 5% and 95% values, and the horizontal line signifies the median. SAS, Self-Rating Anxiety Scale; SDS, Self-Rating Depression Scale; MDD, major depressive disorder; BD, bipolar disorder; ED, eating disorders; \**p <* 0.05; \*\**p <*0.01; \*\*\**p < 0*.001.

The items within the two NSSI functional factors decomposed by OPNMF exhibited higher correlations internally (Figure 2B). A strong negative correlation was observed between the two factors (*r =* −0.766, Figure S1B). Correlation analysis (Figure 2C) revealed influences of depressive and anxious emotions on the two functional factor-loadings. Although SDS and SAS scores were highly correlated among participants (*r =* 0.776, *p* < 0.001), the two factors effectively captured distinct internal characteristics of the patients. Specifically, the self-related factor-loading showed a significant positive correlation with SDS scores (*r =* 0.167, *p* = 0.004), but no significant correlation with SAS scores (*r =* 0.075, *p* = 0.194). However, the social-related factor loading was significantly correlated with SAS scores (*r =* 0.229, *p* < 0.001) but not with SDS scores (*r =* 0.075, *p* = 0.191). Further analysis based on disease diagnosis revealed that the self-related factor-loading was higher in mood disorders (MDD = BD > ED), whereas the social-related factor showed higher loadings in eating disorders (ED > MDD = BD, Figure 2C).

### 3.3 Subtypes of NSSI Function

Fuzzy C-means clustering applied to the two-factor loadings revealed the optimal solution was to divide patients into two clusters (Figure S2). The core members of each cluster were selected based on their likelihoods derived from the fuzzy C-means algorithm, with a cutoff threshold of 0.80. Consequently, two core subtypes of NSSI functions were defined, with 52 ambiguous patients located near the intersection of the clusters (Figure 3A). 意味这目前不以任何功能为主导, 定义为…

**Figure 3.**
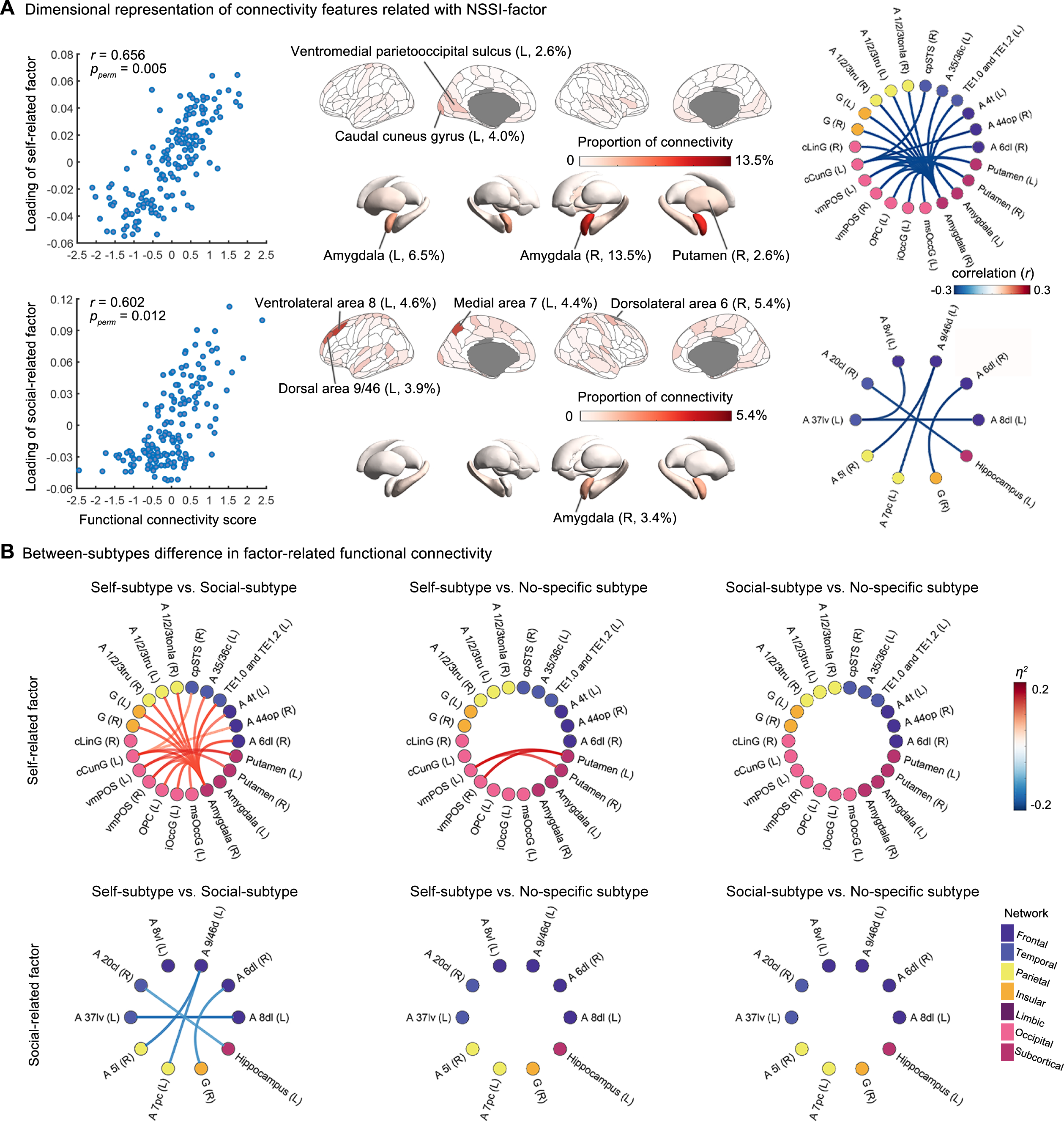
Three NSSI subtypes identified via clustering based on the self–social motivational structure. **(A)** the scatter plot presents a two-dimensional visualization of subtype clustering results. Red points represent social-subtype (social-related subtype, N = 98), gray points represent self-subtype (self-related subtype, N = 154), and blue points represent non-specific subtype (N = 52). **(B)** Violin plots depict the comparison results of the two-factor loadings, depression, and anxiety levels across the three subtypes. **(C)** The bubble chart shows a comparison of risk factors for NSSI among the three subtypes, incorporating six questionnaires: the NEO Five-Factor Inventory (NEOFFI), the revised Chinese Internet Addiction Scale (CIAS), the Adolescent Self-Rating Life Events Check List (ASLEC), the Childhood Trauma Questionnaire (CTQ), the Family Assessment Device (FAD), and the Social Support Rate Scale (SSRS). The horizontal dotted line denotes the significance threshold (*p* < 0.05, FDR corrected), with larger bubbles indicating more significant differences between the groups. **(D)** Risk factors differences between three functional subtypes (post-hoc Turkey test, FDR corrected). Violin plots represent means (dot) and 95% confidence intervals (line). \**p <* 0.05; \*\**p < 0*.01; \*\*\**p < 0*.001.

The largest group included 154 patients (50.7%) and was characterized by higher loadings on the self-related factor and lower loadings on the social-related factor, leading to its designation as the “self-subtype” (Figure 3A). The “social-subtype” comprised 98 patients (32.2%) and was characterized by higher loadings on the social-related factor and lower loadings on the self-related factor. A total of 52 ambiguous patients (17.1%), dispersed between the other two subtypes, were categorized as “non-specific subtype”. No significant differences were found in gender, age, or diagnostic distribution across the three subtypes. Additionally, the subtypes showed similar profiles regarding suicidal ideation, suicide attempts, age of onset, and NSSI frequency/ severity (Table 2 and Table S2).

**Table 2.** Characteristics and functions of NSSI between three functional subtypes.

| | Self-subtype | Social-subtype | Non-specific subtype | $F/\chi^2$ | $p$ |
| --- | --- | --- | --- | --- | --- |
|  | (N = 154,<br>50.7%) | (N = 98,<br>32.2%) | (N = 52,<br>17.1%) |  |  |
| Sex (F/M) | 31/123 | 18/80 | 12/40 | 0.471<sup>a</sup> | 0.790 |
| Age (year) | 18.70 ± 2.46 | 18.36 ± 2.25 | 18.69 ± 2.49 | 0.004 | 0.996 |
| Diagnosis (MDD/BD/ED) | 88/47/19 | 53/26/19 | 26/16/10 | 3.109<sup>a</sup> | 0.540 |
| Suicidal Ideation (Y/N) | 122/32 | 78/20 | 44/8 | 0.755<sup>a</sup> | 0.685 |
| Suicidal Attempt (Y/N) | 90/64 | 62/36 | 27/25 | 1.83<sup>a</sup> | 0.400 |
| NSSI Frequency (R/O) | 92/62 | 57/41 | 29/23 | 0.262<sup>a</sup> | 0.877 |
| Age of onset | 15.31 ± 3.33 | 15.30 ± 3.39 | 14.48 ± 2.36 | 1.329 | 0.266 |
| Self-related factor scores | 29.58 ± 11.73 | 29.22 ± 12.32 | 23.75 ± 15.06 | 4.614 | 0.010<sup>*</sup> |
| Self-related factor loading | 0.07 ± 0.02 | 0.01 ± 0.01 | 0.03 ± 0.02 | 329.324 | <0.001<sup>***</sup> |
| Social-related factor scores | 7.88 ± 5.82 | 23.40 ± 9.58 | 13.40 ± 7.44 | 207.829 | <0.001<sup>***</sup> |
| Social-related factor loading | 0.01 ± 0.01 | 0.08 ± 0.03 | 0.04 ± 0.02 | 557.919 | <0.001<sup>***</sup> |
| Functional scores at onset | 37.51 ± 16.26 | 52.92 ± 20.88 | 37.25 ± 21.72 | 35.553 | <0.001<sup>***</sup> |
| SAS | 59.81 ± 14.70 | 63.43 ± 14.78 | 55.75 ± 12.99 | 4.952 | 0.008<sup>**</sup> |
| SDS | 71.27 ± 13.41 | 71.68 ± 13.93 | 66.40 ± 12.59 | 3.047 | 0.049<sup>*</sup> |
| ASLEC | 46.28 ± 20.02 | 61.51 ± 23.55 | 44.09 ± 21.28 | 18.227 | <0.001<sup>***</sup> |
| FAD | 148.60 ± 23.23 | 147.89 ± 22.19 | 145.09 ± 19.18 | 0.486 | 0.616 |
| CTQ | 47.92 ± 15.13 | 49.84 ± 14.22 | 46.19 ± 14.59 | 1.112 | 0.330 |
| SSRS | 28.84 ± 6.44 | 29.84 ± 6.77 | 31.33 ± 6.52 | 2.886 | 0.057 |
| CIAS | 45.64 ± 14.03 | 41.50 ± 9.05 | 40.71 ± 10.15 | 5.212 | 0.006<sup>**</sup> |
| NEOFFI |  |  |  |  |  |
| Neuroticism | 36.81 ± 6.12 | 39.04 ± 5.91 | 35.63 ± 6.55 | 6.355 | 0.002<sup>**</sup> |
| Extraversion | 21.44 ± 6.69 | 22.27 ± 6.99 | 23.26 ± 6.27 | 1.517 | 0.221 |
| Openness to Experience | 33.56 ± 6.91 | 34.64 ± 6.01 | 32.87 ± 4.96 | 1.524 | 0.219 |
| Agreeableness | 33.52 ± 5.34 | 30.81 ± 5.00 | 32.73 ± 4.47 | 8.532 | <0.001*** |
| Conscientiousness | 29.44 ± 7.29 | 28.99 ± 7.18 | 29.02 ± 7.22 | 0.138 | 0.871 |

Note: R/O, Repeated/Occasional; Y/N, Yes/No; M/F, Male/Female; MDD, major depressive disorder; BD, bipolar disorder; ED, eating disorders; SAS, Self-Rating Anxiety Scale; SDS, Self-Rating Depression Scale; ASLEC, Adolescent Self-Rating Life Events Checklist; FAD, Family Assessment Device; CTQ, Childhood Trauma Questionnaire; SSRS, Social Support Rating Scale; CIAS, Chinese Internet Addiction Scale; NEOFFI, NEO Five-Factor Inventory.
\* $p < 0.05$ ; \*\* $p < 0.01$ ; \*\*\* $p < 0.001$ .
<sup>a</sup>The $\chi^2$ was obtained by a chi-square test.

Regarding NSSI function, social-subtype exhibited a significantly higher total function score compared to the other two subtypes (Cohen’s d = 0.97, *p* < 0.001). Interestingly, in contrast to the factor loadings, social-subtype showed higher scores on both self-related and social-related OSI-F items (Table 2), with multiple functions being reported (Cohen’s d = 0.96, *p* < 0.001). These differences were not significant between self-subtype and non-specific subtype.

Levels of anxiety and depression were significantly different across three subtypes (Figure 3B and Table 2; SAS: Cohen’s d = 0.84, *p* = 0.008; SDS: Cohen’s d = 0.78, *p* = 0.049). Specifically, SAS scores in social-subtype were significantly higher than those in non-specific subtype (Cohen’s d = 0.54, *p* = 0.02); SDS scores in self-subtype and social-subtype were significantly higher compared to non-specific subtype (self vs non-specific: Cohen’s d = 0.37, *p* = 0.024, social vs non-specific: Cohen’s d = 0.39, *p* = 0.023).

Regarding potential NSSI risk factors, no significant differences were found in the total scores for the FAD, CTQ, and SSRS, which assess family functioning, childhood trauma, and social support, respectively, among the three subtypes. Differences were primarily observed in ASLEC, NEOFFI, and CIAS (Figure 3D and Table 2). Specifically, social-subtype reported significantly higher scores on several dimensions of the ASLEC (Cohen’s d = 0.95, *p* < 0.001), including “catch hell,” “loss,” “interpersonal pressure,” “learning pressure,” and “healthy adaptation” (*p* < 0.05, FDR corrected), suggested a higher level of exposure to life events. Similarly, social-subtype exhibited significantly higher levels of internet addiction, as measured by the CIAS (Cohen’s d = 0.85, *p* = 0.006). In terms of personality traits assessed with NEOFFI, social-subtype scored significantly higher in neuroticism compared to the other subtypes (Cohen’s d = 0.87, *p* = 0.002) and significantly lower in agreeableness (Cohen’s d = 0.90, *p* < 0.001).

### 3.4 rsFC Features Related to Functional Factors and Subtypes

We conducted RCCA to investigate the rsFC features that best related to the two-factors of NSSI function using MRI data from 163 participants. To minimize overfitting, we performed a subsampling and cross-validated procedure (see Supplement). This procedure identified two dimensions of rsFC features related to NSSI functions.

The first dimension was significantly positively correlated with loading of self-related factor (Figure 4A, *r =* 0.656, *p =* 0.005), while the second dimension was associated with the loading of social-related factor (*r =* 0.602, *p =* 0.012). We then measured the regional contributions of each dimension (Figure 4A). The top five representative brain regions for the first dimension included the right amygdala (13.5%), left amygdala (6.5%), left caudal cuneus gyrus (4.0%), left ventromedial parietooccipital sulcus (2.6%) and right putamen (2.6%). For the social-related dimension, the top representative brain regions are in the right dorsolateral frontal cortex (Brodmann area, [BA] 6, 5.4%), left ventrolateral frontal cortex (BA8, 4.6%), left medial parietal area (BA7, 4.4%), left dorsal frontal area (BA9/46, 3.9%) and right amygdala (3.4%).

**Figure 4.**
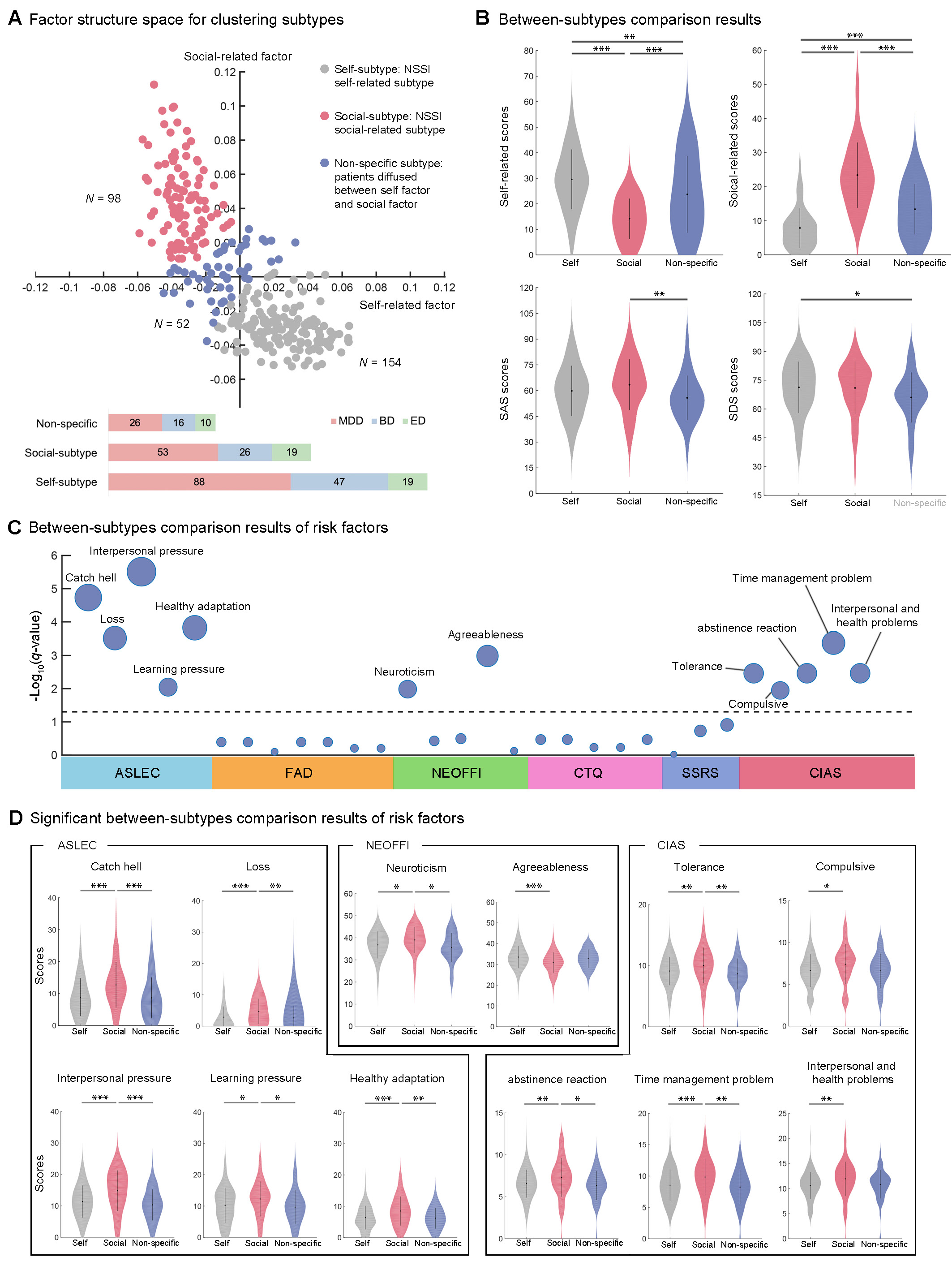
Functional connectivity features correlating with functional factors and subtypes. **(A)** RCC revealed two dimensions related to two functional factors (top: self-related factor; bottom: social-related factor). Scatterplots illustrate the association between connectivity scores and the loading of functional factor scores for each RCC dimension across participants. The schematic diagram of the brain depicts the spatial distribution of regional proportions for each dimension. For each dimension linked to one functional factor, the rsFCs significantly contributing to it were extracted, and the regional proportions were calculated, with darker colors indicating greater contributions. The circular diagram on the far right illustrates the rsFCs significantly associated with the raw scores of the two factors (*p <* 0.001, FDR corrected). Blue lines indicate negative correlations, while red lines represent positive correlations. Node colors differentiate the macroscopic brain regions: purple represents the frontal lobe, blue-purple the temporal lobe, yellow the parietal lobe, orange the insular lobe, dark red the limbic lobe, pink the occipital lobe, and magenta the subcortical nuclei. **(B)** Between-subtype differences in factor-related functional connectivity (top: rsFC correlated with the self-related factor; bottom: rsFC correlated with the social-related factor). From left to right, self-subtype is compared with social-subtype and non-specific subtype, followed by comparisons between the latter two. Red lines indicate that the t-value of comparison is greater than 0, while blue lines indicate that the *T*-value is less than 0.

We then examined the rsFC correlates of each functional factors and the differences between subtypes. The self-related functions were associated with extensive cortico-subcortical functional connectivity (Figure S6). Specifically, the 23 significant connections (surviving a threshold of *p* < 0.001) were observed between the amygdala and various cortical regions, including the superior frontal gyrus (SFG), precentral gyrus, superior temporal gyrus (STG), parahippocampal gyrus, postcentral gyrus, insular gyrus, and occipital lobe. Additionally, rsFC between the putamen and the occipital lobe, as well as subcortical connections between the putamen and the amygdala, were observed (Figure 4A, Table S6). Regarding these self-related connections, self-subtype showed a significantly higher rsFC compared to social-subtype and non-specific subtype, with no differences between the latter two subtypes (Figure 4B).

In contrast, the social-related factor demonstrated a more localized set of rsFC, with six significant connections identified. These connections involved rsFC between the fronto-parietal and fronto-temporal network, as well as connections between the inferior temporal gyrus and hippocampus, dorsolateral superior frontal gyrus and insular gyrus (Figure 4A, Table S6). Social-subtype exhibited elevated rsFC compared to self-subtype, while no significant differences were observed between social-subtype/self-subtype and non-specific subtype (Figure 4B).

### 3.5 Sensitivity Analysis

We conducted an OPNMF on the revised OSI-F (with 7 items deleted) ^41^ for sensitivity analysis. The results indicated that both the self-related and social-related factors comprised 11 items each, confirming a stable and generalizable two-factor structure (Figure S3A-B). The model maintained good inter-factor independence (Figure S3C, *r =* −0.715, *p* < 0.001) and high internal factor consistency (*r =* 0.968, *p* < 0.001). Further imaging verification found that the two factors were still significantly correlated with the previously obtained rsFC characteristics (Figure S3D; self-related: *r =* 0.689, *p* < 0.001; social-related: *r =* 0.774, *p* < 0.001). Additionally, whether for patients with repeated NSSI behaviors or those with occasional NSSI behaviors, the two-factor structure was found to be optimal. Despite the presence of some potentially confusing individual items, the model consistently upheld strong inter-factor independence and internal factor consistency in both groups (Figures S4-S5). The correlation with rsFC characteristics were also reliably replicated across these patient groups (Figures S4-S5).

## Discussion

This study identified two conceptually meaningful and compact factors underlying the motivations and reasons of NSSI behaviors: self-related functions, associated with emotional regulation and self-perception, and social-related functions, linked to interpersonal influence and social interactions. These findings align with the previously proposed intrapersonal and interpersonal functional models of NSSI ^24,42^, providing further empirical support for the heterogeneity of NSSI behaviors. The self-related factor emphasizes on reinforcement from the self, including affect regulation, self-punishment, anti-dissociation, anti-suicide, and marking distress. In contrast, the social-related factor focuses on reinforcement from others or the environment, involving interpersonal influence, establishing boundaries and seeking peer bonding. Importantly, the identified subtypes revealed that self-related and social-related functions are not mutually exclusive but instead represent overlapping motivations, with varying emphases across individuals. While both subtypes exhibited self-related motivations, the social-subtype also demonstrated significant interpersonal concerns, suggesting that self-related motivations are the dominant driver of NSSI behaviors.

Psychosocial factors further differentiate these subtypes. Individuals in the social-subtype reported greater exposure to adverse life events, higher tendency toward problematic internet use, higher neuroticism and lower agreeableness. These findings align with studies showing that social distress can exacerbate emotional dysregulation, reinforcing both self- and social-related motivations for NSSI ^43,44^. Additionally, higher neuroticism was linked to emotional dysregulation and vulnerability to mental health disorders ^45^, while lower agreeableness is associated with difficulties in interpersonal relationships ^46^. These findings suggest offer a streamlined, diagnosis-independent intervention framework for NSSI: emotion regulation strategies may benefit all NSSI subtypes, while those in the social-subtype may additionally require to improve interpersonal effectiveness. Such tailored approaches could address the distinct psychosocial profiles associated with each subtype, potentially enhancing treatment outcomes.

Although the diagnostic distribution of mental disorders did not show significant differences across three subtypes, we observed a notable pattern in the factor loadings. Specifically, the self-related factor loading was higher in mood disorders (MDD and BD), while the social-related factor loading was higher in eating disorders. This suggests that patients with eating disorders may be more likely to engage in NSSI for social-related reasons, such as managing interpersonal relationships or coping with social pressures ^47,48^. Given the relatively small sample size of eating disorder patients in this study, it is important to replicate these findings in larger cohorts to further explore the potential distinctiveness of social-related NSSI motivations in this population.

Neuroimaging analyses revealed distinct neural correlates for self-related and social-related functions. Key findings revealed the amygdala as the central hub for self-related motivations, consistent with its well-documented roles in emotion processing and regulation ^49,50^. Three observed amygdala-centered network (frontal, temporal and insula lobe) constitutes a core substrate for self-related functions. The fronto-amygdala circuitry forms a top-down neural circuit crucial for emotion regulation ^49,51,52^, where enhanced connectivity may paradoxically increase rumination and worry ^53,54^. The temporal-amygdala connectivity supports emotional memory ^55–57^ and processing of affectively charged information ^58^, potentially underlying self-punitive and distress-marking functions. Connections with temporal and insular lobes are involved in self-referential processing ^59^ and may facilitate the transmission of negative emotions, contributing to the emotional dysregulation characteristic of NSSI populations^60,61^. The insula is central to interoception, emotional self-awareness, decision-making, and cognitive control ^62–64^. It has been linked to attention modulation and pain perception in NSSI patients ^65^, as well as deficits in interoceptive awareness and suicidal ideation ^66^. These networks are integral to emotion regulation, self-referential processing, and interoceptive awareness, supporting the emotional and self-punitive motivations underlying self-subtype of NSSI.

In contrast, social-related functions were linked to fronto-parietal, fronto-insular, and fronto-temporal networks, which are involved in social cognition, interpersonal behavior, and the regulation of social-emotional information. The dorsal fronto-parietal network supports top-down attentional orienting processes ^67,68^ and is recruited during the relational integration of social information ^69^. The medial prefrontal cortex and precuneus (the medial portion of BA7) within this network play pivotal roles in adaptive interpersonal behavior ^70^ and self-related mental representations ^71,72^. The insula exhibits consistent activation under social exclusion ^73^. The fronto-insula hyperconnectivity appears linked to maladaptive introspection ^74^ and negative emotional information ^75^. Concurrently, the fronto-temporal network also participates in the “social brain”, and within this network, fusiform gyrus has been implicated in socioemotional processes ^76^ interpersonal influence ^71^.These neural circuits reflect the higher rejection sensitivity combined with ineffective attempts to regulate feelings of rejection in individuals with NSSI, underscoring their difficulties in emotion regulation within social contexts. Moreover, these findings support the potential translational utility of neuromodulation strategies targeting subtype-specific neural circuits in adolescents with NSSI.

The application of fuzzy clustering allowed us to capture the heterogeneity of NSSI behaviors more effectively ^35^. The identification of an undifferentiated subtype (non-specific subtype), characterized by lower scores on both self- and social-related motivations, suggests the existence of individuals who have yet to develop a dominant function for NSSI. This subgroup highlights the potential for shifts in motivational bases over time, warranting further longitudinal investigation to track changes in NSSI functions and their associated neurobiological profiles.

Despite these advances, several limitations should be acknowledged. Firstly, the cross-sectional design limits our ability to infer causality or assess the temporal stability of the identified functional subtypes. Longitudinal studies are needed to explore the evolution of NSSI motivations and their neural correlates over time, especially for non-specific subtype. Secondly, these results need to be replicated and validated using independent samples. Finally, the relatively small sample size of certain subgroups, such as individuals with eating disorders, may limit the generalizability of our results. Meanwhile, as a cross-culture behavior, future studies with larger, culturally diverse cohorts are needed to further explore the distinctiveness of social-related NSSI expression and motivations.

In conclusion, this study bridges the gap between behavioral functions and neurobiological phenotypes in NSSI, an area that has received limited attention in prior research. By identifying distinct NSSI subtypes with corresponding neural and psychosocial profiles, our findings provide a foundation for developing more precise and effective interventions. Tailoring treatments to the specific functional and neurobiological characteristics of individuals with NSSI may enhance therapeutic outcomes and ultimately reduce the burden of this debilitating behavior.

## Data Availability

All data produced in the present study are available upon reasonable request to the authors.

## Acknowledgements

We extend our gratitude to all participants in this study. This study was supported by the National Natural Science Foundation of China (82071505, 81771443, 81361120395). HY was partly supported by Clinical Medicine Plus X – Young Scholars Project, Peking University the Fundamental Research Funds for the Central Universities (PKU2024LCXQ046) and Capital’s Funds for Health Improvement and Research (2024-2-4115). We also acknowledge the development of a preliminary MATLAB-based toolkit for classifying NSSI subtypes, available at https://github.com/XiaoYangLab/NSSI_factor.

## Reference

1 Nock MK. Self-Injury. Annu Rev Clin Psychol 2010; 6: 339–363.

2 Singhal A, Ross J, Seminog O, Hawton K, Goldacre MJ. Risk of self-harm and suicide in people with specific psychiatric and physical disorders: comparisons between disorders using English national record linkage. J R Soc Med 2014; 107: 194–204.

3 Swannell SV, Martin GE, Page A, Hasking P, St John NJ. Prevalence of Nonsuicidal Self-Injury in Nonclinical Samples: Systematic Review, Meta-Analysis and Meta-Regression. Suicide Life Threat Behav 2014; 44: 273–303.

4 Wolff J, Frazier EA, Esposito-Smythers C, Burke T, Sloan E, Spirito A. Cognitive and Social Factors Associated with NSSI and Suicide Attempts in Psychiatrically Hospitalized Adolescents. Journal of Abnormal Child Psychology 2013; 41: 1005–1013.

5 Wang L, Liu J, Yang Y, Zou H. Prevalence and risk factors for non-suicidal self-injury among patients with depression or bipolar disorder in China. BMC Psychiatry 2021; 21: 389.

6 Ribeiro JD, Franklin JC, Fox KR, Bentley KH, Kleiman EM, Chang BP et al. Self-injurious thoughts and behaviors as risk factors for future suicide ideation, attempts, and death: a meta-analysis of longitudinal studies. Psychol Med 2016; 46: 225–236.

7 Tan AC, Rehfuss MC, Suarez EC, Parks-Savage A. Nonsuicidal self-injury in an adolescent population in Singapore. Clin Child Psychol Psychiatry 2014; 19: 58–76.

8 Qu D, Wen X, Liu B, Zhang X, He Y, Chen D et al. Non-suicidal self-injury in Chinese population: a scoping review of prevalence, method, risk factors and preventive interventions. The Lancet Regional Health–Western Pacific 2023; 37.

9 Qu D, Wen X, Liu B, Zhang X, He Y, Chen D et al. Non-suicidal self-injury in Chinese population: a scoping review of prevalence, method, risk factors and preventive interventions. Lancet Reg Health West Pac 2023; 37: 100794.

10 Wang Y-J, Li X, Ng CH, Xu D-W, Hu S, Yuan T-F. Risk factors for non-suicidal self-injury (NSSI) in adolescents: A meta-analysis. EClinicalMedicine 2022; 46: 101350.

11 Wolff JC, Thompson E, Thomas SA, Nesi J, Bettis AH, Ransford B et al. Emotion dysregulation and non-suicidal self-injury: A systematic review and meta-analysis. Eur Psychiatry 2019; 59: 25–36.

12 Tatnell R, Kelada L, Hasking P, Martin G. Longitudinal Analysis of Adolescent NSSI: The Role of Intrapersonal and Interpersonal Factors. Journal of Abnormal Child Psychology 2014; 42: 885–896.

13 Calvo N, García-González S, Perez-Galbarro C, Regales-Peco C, Lugo-Marin J, Ramos-Quiroga J-A et al. Psychotherapeutic interventions specifically developed for NSSI in adolescence: A systematic review. Eur Neuropsychopharmacol 2022; 58: 86–98.

14 Jiang Z, Wang Z, Diao Q, Chen J, Tian G, Cheng X et al. Efficacy of interventions for suicide and self-injury in children and adolescents: a meta-analysis. Sci Rep 2023; 12: 12313.

15 Kaess M, Hooley JM, Klimes-Dougan B, Koenig J, Plener PL, Reichl C et al. Advancing a temporal framework for understanding the biology of nonsuicidal self-injury: An expert review. Neuroscience & Biobehavioral Reviews 2021; 130: 228–239.

16 Whittle S, Zhang L, Rakesh D. Environmental and neurodevelopmental contributors to youth mental illness. Neuropsychopharmacology 2024; 50: 201–210.

17 Auerbach RP, Pagliaccio D, Allison GO, Alqueza KL, Alonso MF. Neural Correlates Associated With Suicide and Nonsuicidal Self-injury in Youth. Biological Psychiatry 2021; 89: 119–133.

18 Brañas MJAA, Croci MS, Ravagnani Salto AB, Doretto VF, Martinho E, Macedo M et al. Neuroimaging Studies of Nonsuicidal Self-Injury in Youth: A Systematic Review. Life 2021; 11: 729.

19 Morawetz C, Riedel MC, Salo T, Berboth S, Eickhoff SB, Laird AR et al. Multiple large-scale neural networks underlying emotion regulation. Neuroscience & Biobehavioral Reviews 2020; 116: 382–395.

20 Ho TC, Walker JC, Teresi GI, Kulla A, Kirshenbaum JS, Gifuni AJ et al. Default mode and salience network alterations in suicidal and non-suicidal self-injurious thoughts and behaviors in adolescents with depression. Transl Psychiatry 2021; 11: 38.

21 Liu X, Hairston J, Schrier M, Fan J. Common and distinct networks underlying reward valence and processing stages: a meta-analysis of functional neuroimaging studies. Neurosci Biobehav Rev 2011; 35: 1219–1236.

22 Wang Z, Li D, Chen Y, Tao Z, Jiang L, He X et al. Understanding the subtypes of non-suicidal self-injury: A new conceptual framework based on a systematic review. Psychiatry Research 2024; 334: 115816.

23 Nock MK, Prinstein MJ, Nock MK, Prinstein MJ. A Functional Approach to the Assessment of Self-Mutilative Behavior. Journal of Consulting and Clinical Psychology 2004; 72: 885–890.

24 Klonsky ED, Glenn CR, Styer DM, Olino TM, Washburn JJ. The functions of nonsuicidal self-injury: converging evidence for a two-factor structure. Child Adolesc Psychiatry Ment Health 2015; 9: 44.

25 Taylor PJ, Jomar K, Dhingra K, Forrester R, Shahmalak U, Dickson JM. A meta-analysis of the prevalence of different functions of non-suicidal self-injury. Journal of Affective Disorders 2018; 227: 759–769.

26 Nock MK, Prinstein MJ. Contextual features and behavioral functions of self-mutilation among adolescents. Journal of abnormal psychology 2005; 114: 140.

27 Park Y, Qu W, Ammerman BA. Characteristics and Functions of Non-Suicidal Self-Injury That Inform Suicide Risk. Archives of Suicide Research 2024; : 1–14.

28 [CHENG Wen-hong ZF, et al, XIAO Ze-ping. Study on reliability and validity of Chinese version of Ottawa self-injury inventory. Journal of Shanghai Jiao Tong University (Medical Science) 2015; 35: 460-.

29 Xiang Dong Wang, Wang XL, Ma H. Rating scales for mental health. Beijing: Chinese Mental Health Journal Press, 1999.

30 Huang XT. Handbook of personality: theory and research. Shanghai: East China Normal University Press, doi 2003; 10: 2093310.

31 Bai Y, Fan F. A Study on the Internet Dependence of College Students: the Revising and Applying of a Measurement. Psychological Development and Education 2005; 21: 99–104.

32 Zhao X, Zhang Y, Li L, Zhou Y. Evaluation on reliability and validity of Chinese version of childhood trauma questionnaire. Chinese Journal of Tissue Engineering Research 2005; : 209–211.

33 Sotiras A, Resnick SM, Davatzikos C. Finding imaging patterns of structural covariance via Non-Negative Matrix Factorization. NeuroImage 2015; 108: 1–16.

34 Zhirong Yang, Oja E. Linear and Nonlinear Projective Nonnegative Matrix Factorization. IEEE Trans Neural Netw 2010; 21: 734–749.

35 Chen J, Patil KR, Weis S, Sim K, Nickl-Jockschat T, Zhou J et al. Neurobiological Divergence of the Positive and Negative Schizophrenia Subtypes Identified on a New Factor Structure of Psychopathology Using Non-negative Factorization: An International Machine Learning Study. Biological Psychiatry 2020; 87: 282–293.

36 Bezdek JC. Pattern Recognition With Fuzzy Objective Function Algorithms. Pattern Recognition with Fuzzy Objective Function Algorithms, 1981.

37 Yan CG, Wang XD, Zuo XN, Zang YF. DPABI: Data Processing & Analysis for (Resting-State) Brain Imaging. Neuroinformatics 2016; 14: 339–351.

38 Fan L, Li H, Zhuo J, Zhang Y, Wang J, Chen L et al. The Human Brainnetome Atlas: A New Brain Atlas Based on Connectional Architecture. Cereb Cortex 2016; 26: 3508–3526.

39 Desikan RS, Ségonne F, Fischl B, Quinn BT, Dickerson BC, Blacker D et al. An automated labeling system for subdividing the human cerebral cortex on MRI scans into gyral based regions of interest. Neuroimage 2006; 31: 968–980.

40 Buch AM, Vértes PE, Seidlitz J, Kim SH, Grosenick L, Liston C. Molecular and network-level mechanisms explaining individual differences in autism spectrum disorder. Nat Neurosci 2023; 26: 650–663.

41 Chen H, Pan B, Zhang C, Guo Y, Zhou J, Wang X. Revision of the non-suicidal self-injury behavior scale for adolescents with mental disorder. Zhong Nan Da Xue Xue Bao Yi Xue Ban 2022; 47: 301–308.

42 Klonsky ED, Glenn CR. Assessing the Functions of Non-suicidal Self-injury: Psychometric Properties of the Inventory of Statements About Self-injury (ISAS). J Psychopathol Behav Assess 2009; 31: 215–219.

43 Tang J, Ma Y, Lewis SP, Chen R, Clifford A, Ammerman BA et al. Association of Internet Addiction With Nonsuicidal Self-injury Among Adolescents in China. JAMA Network Open 2020; 3: e206863–e206863.

44 Yen J-Y, Yen C-F, Chen C-S, Wang P-W, Chang Y-H, Ko C-H. Social anxiety in online and real-life interaction and their associated factors. Cyberpsychol Behav Soc Netw 2012; 15: 7–12.

45 Ormel J, Jeronimus BF, Kotov R, Riese H, Bos EH, Hankin B et al. Neuroticism and common mental disorders: meaning and utility of a complex relationship. Clin Psychol Rev 2013; 33: 686–697.

46 Graziano WG, Jensen-Campbell LA, Hair EC. Perceiving interpersonal conflict and reacting to it: the case for agreeableness. J Pers Soc Psychol 1996; 70: 820–835.

47 Muehlenkamp JJ, Takakuni S, Brausch AM, Peyerl N. Behavioral functions underlying NSSI and eating disorder behaviors. J Clin Psychol 2019; 75: 1219–1232.

48 Kiekens G, Claes L. Non-Suicidal Self-Injury and Eating Disordered Behaviors: An Update on What We Do and Do Not Know. Curr Psychiatry Rep 2020; 22: 68.

49 Morawetz C, Riedel MC, Salo T, Berboth S, Eickhoff SB, Laird AR et al. Multiple large-scale neural networks underlying emotion regulation. Neuroscience & Biobehavioral Reviews 2020; 116: 382–395.

50 Ochsner KN, Silvers JA, Buhle JT. Functional imaging studies of emotion regulation: a synthetic review and evolving model of the cognitive control of emotion. Ann N Y Acad Sci 2012; 1251: E1–24.

51 Ochsner KN, Bunge SA, Gross JJ, Gabrieli JD. Rethinking feelings: an FMRI study of the cognitive regulation of emotion. Journal of cognitive neuroscience 2002; 14: 1215–1229.

52 Refojo D, Schweizer M, Kuehne C, Ehrenberg S, Thoeringer C, Vogl AM et al. Glutamatergic and dopaminergic neurons mediate anxiogenic and anxiolytic effects of CRHR1. Science 2011; 333: 1903–1907.

53 Murphy ER, Barch DM, Pagliaccio D, Luby JL, Belden AC. Functional connectivity of the amygdala and subgenual cingulate during cognitive reappraisal of emotions in children with MDD history is associated with rumination. Dev Cogn Neurosci 2016; 18: 89–100.

54 Sokołowski A, Kowalski J, Dragan M. Neural functional connectivity during rumination in individuals with adverse childhood experiences. Eur J Psychotraumatol 2022; 13: 2057700.

55 LaBar KS, Cabeza R. Cognitive neuroscience of emotional memory. Nat Rev Neurosci 2006; 7: 54–64.

56 Smith APR, Stephan KE, Rugg MD, Dolan RJ. Task and content modulate amygdala-hippocampal connectivity in emotional retrieval. Neuron 2006; 49: 631–638.

57 Kark SM, Slotnick SD, Kensinger EA. Repetition Enhancement of Amygdala and Visual Cortex Functional Connectivity Reflects Nonconscious Memory for Negative Visual Stimuli. J Cogn Neurosci 2016; 28: 1933–1946.

58 LeDoux JE. Emotion Circuits in the Brain.

59 van der Meer L, Costafreda S, Aleman A, David AS. Self-reflection and the brain: a theoretical review and meta-analysis of neuroimaging studies with implications for schizophrenia. Neurosci Biobehav Rev 2010; 34: 935–946.

60 Nam G, Moon H, Lee J-H, Hur J-W. Self-referential processing in individuals with nonsuicidal self-injury: An fMRI study. Neuroimage Clin 2022; 35: 103058.

61 Liao K, Yu R, Chen Y, Chen X, Wu X, Huang X et al. Alterations of regional brain activity and corresponding brain circuits in drug-naïve adolescents with nonsuicidal self-injury. Sci Rep 2024; 14: 24997.

62 Farb NAS, Segal ZV, Anderson AK. Attentional modulation of primary interoceptive and exteroceptive cortices. Cereb Cortex 2013; 23: 114–126.

63 Craig ADB. How do you feel--now? The anterior insula and human awareness. Nat Rev Neurosci 2009; 10: 59–70.

64 Critchley HD. Neural mechanisms of autonomic, affective, and cognitive integration. J Comp Neurol 2005; 493: 154–166.

65 Krause-Utz A, Frost R, Winter D, Elzinga BM. Dissociation and Alterations in Brain Function and Structure: Implications for Borderline Personality Disorder. Curr Psychiatry Rep 2017; 19: 1–22.

66 Ho TC, Walker JC, Teresi GI, Kulla A, Kirshenbaum JS, Gifuni AJ et al. Default mode and salience network alterations in suicidal and non-suicidal self-injurious thoughts and behaviors in adolescents with depression. Transl Psychiatry 2021; 11: 38.

67 Petersen SE, Posner MI. The attention system of the human brain: 20 years after. Annu Rev Neurosci 2012; 35: 73–89.

68 Corbetta M, Shulman GL. Control of goal-directed and stimulus-driven attention in the brain. Nat Rev Neurosci 2002; 3: 201–215.

69 Magis-Weinberg L, Blakemore S-J, Dumontheil I. Social and Nonsocial Relational Reasoning in Adolescence and Adulthood. J Cogn Neurosci 2017; 29: 1739–1754.

70 Bickart KC, Dickerson BC, Feldman Barrett L. The amygdala as a hub in brain networks that support social life. Neuropsychologia 2014; 63: 235–248.

71 Quevedo K, Martin J, Scott H, Smyda G, Pfeifer JH. The neurobiology of self-knowledge in depressed and self-injurious youth. Psychiatry Res Neuroimaging 2016; 254: 145–155.

72 Cavanna AE, Trimble MR. The precuneus: a review of its functional anatomy and behavioural correlates. Brain 2006; 129: 564–583.

73 Cacioppo S, Frum C, Asp E, Weiss RM, Lewis JW, Cacioppo JT. A Quantitative Meta-Analysis of Functional Imaging Studies of Social Rejection. Scientific Reports 2013; 3: 2027.

74 Kaiser RH, Whitfield-Gabrieli S, Dillon DG, Goer F, Beltzer M, Minkel J et al. Dynamic Resting-State Functional Connectivity in Major Depression. Neuropsychopharmacology 2016; 41: 1822–1830.

75 Kaiser RH, Snyder HR, Goer F, Clegg R, Ironside M, Pizzagalli DA. Attention Bias in Rumination and Depression: Cognitive Mechanisms and Brain Networks. Clin Psychol Sci 2018; 6: 765–782.

76 Suarez GL, Burt SA, Gard AM, Burton J, Clark DA, Klump KL et al. The impact of neighborhood disadvantage on amygdala reactivity: Pathways through neighborhood social processes. Dev Cogn Neurosci 2022; 54: 101061.

