## Supplemental Materials for "Functional Heterogeneity in Non-Suicidal Self-Injury across Psychiatric Disorders: Neural and Psychosocial Correlates"

### SUPPLEMENTARY MATERIALS

### I. Supplementary Methods

#### 1.1 Model evaluation for selecting the optimal number of factors in OPNMF

To determine the most robust and stable factor model as a dimensionality reduced conceptualization of NSSI function, a sophisticated evaluation scheme was performed according to previous study[1]. In this study, we employed a set of sophisticated evaluation strategies based on cross-validation with 10,000 split-half analyses. Specifically, the set of columns of the original OSI-F item by entire patient matrix (M) was randomly split into two submatrices (M1 and M2), which were independent and equal length. OPNMF was conducted separately on the M1 and M2, and each item in OSI-F was assigned to a certain factor for M1/M2. Then, we used some evaluation indicators to evaluate the stability and generalizability of the factor model.

Three evaluation indicators were computed to assess model stability. By hard-assigning items to specific factors (as a form of natural clustering), the adjusted Rand index (RI)[2] and variation of information (VI)[3] were utilized to indicate the similarity of factor-label assignment for each item and item-pair grouping between the two split samples in each submatrix (M1 and M2). Higher values for adjusted RI and lower values for VI denote greater stability. Additionally, considering that one item can be influenced by multiple dimensions and may have minor contributions to other factors (low coefficients loaded on other factors aside from the one to which an item is assigned), the concordance index (CI)[4], which reflect the concordance of cosine similarity for each pair of OSF-F items between the factorizations of split-samples (M1 and M2), was employed to take into account items with multiple factor-memberships. Higher values for CI denote greater model stability.

The generalizability of the factor model was assessed by measuring the increase in out-of-sample reconstruction error (RE)[1]. The RE is defined as the absolute difference between the reconstructed matrix and the original data matrix. An increase in out-of-sample RE reflects the degree to which the matrix reconstruction is worse when using the dictionary (basis matrix) obtained from a model-unseen sample, compared to the reconstruction error calculated from the within-sample dictionary. A smaller increase in out-of-sample RE, relative to the within-sample reconstruction error, suggests superior generalizability.

**1.2 Evaluation for the optimal number of the fuzzy C-means clustering**

Conventional clustering methods, such as k-means, operate under a "hard" clustering framework, where each individual is assigned to a single, mutually exclusive cluster. This approach assumes clear-cut category boundaries, which may not accurately reflect the complex and overlapping nature of psychopathological traits.

In contrast, Fuzzy C-means clustering is an unsupervised machine learning algorithm that extends the traditional k-means method by allowing each subject to belong to multiple clusters simultaneously, with a degree of membership ranging from 0 to 1. This probabilistic assignment enables the modeling of ambiguous or transitional profiles, which is especially relevant in psychiatric research where diagnostic boundaries are often fluid. During the clustering process, it is essential to ascertain the optimal number of clusters within a dataset. The cluster validity index was utilized to identify the optimal number of clusters from the dataset[1].

We used the fuzzy silhouette index[5] to determine the optimal number of clusters. This index requires two components: first, the dataset partitioned by the clustering technique (in this case, fuzzy C-means), and second, the calculation of similarities between data vectors, represented by their Euclidean distances. A higher silhouette value indicates better-defined clusters.

The Xie-Beni Index[6] is commonly applied in fuzzy clustering to determine the number of optimal clusters. Specifically, it is calculated as the sum of squared distances between each data point and the centroid of its respective cluster, normalized by the minimum distance between cluster centroids. This index penalizes clusters that are either too diffuse or too close to one another, thereby favoring more compact and well-separated clusters. The smaller values of Xie-Beni Index represent for more compact and well-separated clusters.

Partition Entropy (PE)[7] offers a quantitative assessment regarding how effectively the data is partitioned into clusters, concentrating on the degree of uncertainty or disorder in the cluster assignments. Smaller values of PE signify a more organized and distinct clustering structure, whereas larger values imply less clearly defined clusters and greater ambiguity in the assignment of data points to clusters.

To determine the optimal cutoff for membership discrimination, we applied the elbow method within a range of 70% to 90%. This method is based on the average degree of distortion, measured in terms of squared distances[8]. The elbow point, where the rate of decrease in explained variance slowed significantly, was identified, representing a balance between maximizing cluster compactness and minimizing the number of clusters.

**1.3 MRI scans parameters**

Images data of all participants were acquired on a 3.0 T GE Discovery MR750 scanner in the Center for Neuroimaging, Peking University Sixth Hospital (Clinical dataset). The high-resolution structural T1-weighted MRI was acquired in a sagittal orientation using an axial 3D fast, spoiled gradient recalled (FSPGR) sequence with the following parameters: time repetition (TR) = 2500 ms, time echo (TE) = 2.38 ms, field of view (FOV) = 256 × 256 $mm^2$, slice thickness/gap = 0.8/0 mm, acquisition voxel size = 0.8 × 0.8 × 0.8 $mm^3$, flip angle = 8°, 224 contiguous sagittal slices. For the resting-state functional, the parameters were: TR = 2000 ms, TE = 34.0 ms, FOV = 200 × 200 $mm^2$, flip angle = 90°, voxel size = 2 × 2 × 2 $mm^3$, and 240 slices.

**1.4 Resting-state functional MRI preprocessing and quality control**

Data preprocessing was conducted using the DPABI (Data Processing Assistant for Resting State fMRI Advanced Edition)[9] software based on the MATLAB (The MathWorks, Natick, MA, USA) with a standardized protocol.

Specifically, image processing included 1) removing initial 10 volumes for signal equilibrium and environmental adaptation; 2) performing slice-timing correction in remaining images; 3) realigning remaining images to the first volume for head motion correction using a six-parameter (rigid body) linear transformation; 4) registration and spatial standardization: register the functional image with the T1 structural image. Conduct spatial standardization through Diffeomorphic Anatomical Registration

Through Exponentiated Lie Algebra (DARTEL); 5) performing the spatial smoothing with a Gaussian filter set at 8mm full-width at half-maximum; 6) remove linear drift to eliminate the influence of linear trends; 7) use linear regression to remove the influence of covariates, including head motion (using the Friston 24-parameter model)[10], white matter signal, and cerebrospinal fluid signal; 8) low-pass filtering (0.01 to 0.1 Hz).

After preprocessing, quality control based on head motion correction and spatial standardization is performed. Samples with head motion ≥ 3 (translation exceeding 3 mm or rotation exceeding 3°) are excluded.

**1.5 Details of regularized canonical correlation analysis (RCC)**

To better understand the neural correlates of NSSI functions, we employed a start-of-art association analysis to capture the relationship between NSSI functional factors and rsFC. An extended association analysis combining dimensional representation with feature selection was thought to better capture the monotonic relationship between the clinical features and neuroimaging measurements. Therefore, we trained a machine-learning framework based on regularized canonical correlation (RCC) with a hyper-parameterized feature selection space following a previous study[11] to explore the relationship between factor structures and rsFC (Figure S7).

Specifically, the RCC is a statistical technique widely used to explore the dimensional representations between two sets of variables. It could be used to identify pairs of linear combinations (called canonical variates) such that the correlation between the canonical variates from each set is maximized. By introducing an L2 penalty ('ridge' or 'Tikhonov' penalty), the RCC method could handle multicollinearity in the input data to avoid potential overfitting to some extent. In the current study, firstly, the RCC was embedded into a nested cross-validation. We split the dataset (N = 163) into training data (N = 146) and testing data according to a 90% random selection, with 100 replications. For each replication, the training data was further split into RCC-training data (N = 139) and validation data according to a 90% random selection, with 30 replications, in which the RCC model would be trained based on RCC-training data and tested using validation data. Next, to conduct feature selection, we also take the number of potential rsFC features ($N_{features}$) as a parameter, and put it into the hyperparameter space together with L2-penalty ($\lambda$) to optimization. The $N_{features}$ refers to the number of rsFC selected to train the RCC model from a bootstrapping scheme. This scheme further subsampled the RCC-training data according to a 95% ratio, and ranked rsFC features based on a mean spearman correlation coefficient with factor-loadings of NSSI functions across 100 times subsampling replications. The $N_{features}$ top-ranked features was be exacted to train the RCC model. After a definition of hyperparameter space, we conducted a grid search to settle optimal parameters from a hyperparameter grid of $N_{features}$ on the 100 to 400 top-ranked rsFC and $\lambda$ on the 1 to 10. Benefit from nested cross-validation measurement, all processes of optimization were carried out in the training data, which have been proved could effectively avoid the information leakage in the model training. Then, based on the optimal parameters, we calculated the canonical correlation coefficients between NSSI functional factors and rsFC in testing data (hold-out data) to represent the model performance and the statistical significance of coefficients was evaluated using a 10,000 times nonparametric permutation test. Finally, the procedures yielded canonical correlation coefficients for all latent factors and canonical variate pairs for all participants to represent the factor-rsFC relationship and dimensional representation of rsFC (FC scores derive from RCC model), respectively.

##### 1.6 Sample Size Justification

The total sample in the present study comprised 304 participants with psychiatric disorders who completed the Ottawa Self-Injury Inventory (OSI) for functional factor analysis. The present sample size is consistent with prior studies examining the functional structure of non-suicidal self-injury (NSSI). For example, Klonsky et al.[12]assessed 235 college students using the Inventory of Statements About Self-Injury (ISAS) with exploratory factor analysis, Rodav et al.[13] analyzed 275 high school adolescents using the OSI with exploratory factor analysis, and Case et al.[14] examined 359 undergraduates using the Deliberate Self-Harm Inventory (DSHI) with latent class analysis.

#### II. Supplementary Tables

##### Table S1: NSSI episodes and functions

| | All Samples (N = 304) | MRI Samples (N = 163) | *T/χ2* | *p* |
|---|---|---|---|---|
| **Suicidal Ideation (Yes)** | 244 (80.3%) | 131 (80.4%) | 0.001 [a] | 0.978 |
| **Suicidal Attempt (Yes)** | 177 (58.2%) | 89 (54.6%) | 0.568 [a] | 0.451 |
| **NSSI frequency in past month** | | | 0.28 [a] | 0.964 |
| not at all | 67 (22.0%) | 33 (20.2%) | | |
| at least once | 126 (41.4%) | 71 (43.6%) | | |
| weekly | 74 (24.3%) | 39 (23.9%) | | |
| daily | 37 (12.2%) | 20 (12.3%) | | |
| **NSSI frequency in past 6 months** | | | 0.163 [a] | 0.997 |
| not at all | 25 (8.2%) | 14 (8.6%) | | |
| 1 to 5 times | 159 (52.3%) | 84 (51.5%) | | |
| monthly | 36 (11.8%) | 21 (12.9%) | | |
| weekly | 60 (19.7%) | 32 (19.6%) | | |
| daily | 24 (7.9%) | 12 (7.4%) | | |
| **NSSI frequency in the past year** | | | 0.566 [a] | 0.967 |
| not at all | 28 (9.2%) | 16 (9.8%) | | |
| 1 to 5 times | 153 (50.3%) | 81 (49.7%) | | |
| monthly | 59 (19.4%) | 30 (18.4%) | | |
| weekly | 48 (15.8%) | 29 (17.8%) | | |
| daily | 16 (5.3%) | 7 (4.3%) | | |
| **NSSI frequency in one year ago** | | | 3.177 [a] | 0.529 |
| not at all | 77 (25.3%) | 45 (27.6%) | | |
| 1 to 5 times | 130 (42.8%) | 62 (38.0%) | | |
| monthly | 35 (11.5%) | 21 (12.9%) | | |
| weekly | 43 (14.1%) | 29 (17.8%) | | |
| daily | 19 (6.3%) | 6 (3.7%) | | |
| **Age of onset** | 15.17 ± 3.21 | 15.06 ± 3.17 | 0.322 | 0.748 |
| **NSSI function at onset** | | | | |
| Total functional scores | 42.43 ± 20.12 | 42.00 ± 18.66 | 0.295 | 0.821 |
| Number of functions endorsed | 16.22 ± 6.10 | 16.57 ±5.91 | 0.604 | 0.546 |
| Number of body parts injured | 3.66 ± 2.30 | 3.86 ± 2.36 | 0.892 | 0.373 |
| **NSSI function last month** | | | | |
| Total functional scores | 42.68 ± 22.63 | 41.16 ± 21.02 | 0.626 | 0.532 |
| Number of functions endorsed | 15.78 ± 6.73 | 15.91 ± 6.67 | 0.187 | 0.851 |
| Number of body parts injured | 2.61 ± 1.80 | 2.70 ± 1.86 | 0.603 | 0.666 |

[a] The $\chi^2$ was obtained by a chi-square test.

#### Table S2: NSSI episodes and functions of three subtypes

| | Self-subtype (N = 154, 50.7%) | Social-subtype (N = 98, 32.2%) | Non-specific subtype (N = 52, 17.1%) | *F/χ2* | *p* |
|---|---|---|---|---|---|
| **Frequency (Repeated/ Occasional)** | 92/62 | 57/41 | 29/23 | 0.262 [a] | 0.877 |
| **Self-related factor scores** | 29.58 ± 11.73 | 29.22 ± 12.32 | 23.75 ± 15.06 | 4.461 | 0.012* |
| **Self-related factor loading** | 0.07 ± 0.02 | 0.01 ± 0.01 | 0.03 ± 0.02 | 329.324 | <0.001*** |
| **Social-related factor scores** | 7.88 ± 5.82 | 23.40 ± 9.58 | 13.40 ± 7.44 | 128.48 | <0.001*** |
| **Social-related factor loading** | 0.01 ± 0.01 | 0.08 ± 0.03 | 0.04 ± 0.02 | 557.919 | <0.001*** |
| **Suicidal Ideation (Yes)** | 122 (79.2%) | 78 (79.6%) | 44 (84.6%) | 0.755 [a] | 0.685 |
| **Suicidal Attempt (Yes)** | 90 (58.4) | 62 (63.3%) | 27 (51.9%) | 1.83 [a] | 0.400 |
| **NSSI frequency in past month** | | | | 4.529 [a] | 0.606 |
| not at all | 39 (25.3%) | 19 (19.4%) | 9 (17.3%) | | |
| at least once | 57 (37.0%) | 43 (43.9%) | 26 (50.0%) | | |
| weekly | 37 (24.0%) | 24 (24.5%) | 13 (25.0%) | | |
| daily | 21 (13.6%) | 12 (12.2%) | 4 (7.7%) | | |
| **NSSI frequency in past 6 months** | | | | 5.229 [a] | 0.733 |
| not at all | 16 (10.4%) | 7 (7.1%) | 2 (3.8%) | | |
| 1 to 5 times | 74 (48.1%) | 55 (56.1%) | 30 (57.7%) | | |
| monthly | 19 (12.3%) | 9 (9.2%) | 8 (15.4%) | | |
| weekly | 32 (20.8%) | 20 (20.4%) | 8 (15.4%) | | |
| daily | 13 (8.4%) | 7 (7.1%) | 4 (7.7%) | | |
| **NSSI frequency in the past year** | | | | 6.668 [a] | 0.573 |
| not at all | 14 (9.1%) | 10 (10.2%) | 4 (7.7%) | | |
| 1 to 5 times | 77 (50.0%) | 47 (48.0%) | 29 (55.8%) | | |
| monthly | 28 (18.2%) | 20 (20.4%) | 11 (21.2%) | | |
| weekly | 30 (19.5%) | 13 (13.3%) | 5 (9.6%) | | |
| daily | 5 (3.2%) | 8 (8.2%) | 3 (5.8%) | | |
| **NSSI frequency in one year ago** | | | | 4.821 [a] | 0.776 |
| not at all | 42 (27.3%) | 21 (21.4%) | 14 (26.9%) | | |

| | | | | | |
|---|---|---|---|---|---|
| 1 to 5 times | 62 (40.3%) | 48 (49.0%) | 20 (38.5%) | | |
| monthly | 19 (12.3%) | 8 (8.2%) | 8 (15.4%) | | |
| weekly | 23 (14.9%) | 14 (14.3%) | 6 (11.5%) | | |
| daily | 8 (5.2%) | 7 (7.1%) | 4 (7.7%) | | |
| **Age of onset** | 15.31 ± 3.33 | 15.30 ± 3.39 | 14.48 ± 2.36 | 1.329 | 0.266 |
| **At onset** | | | | | |
| Total functional scores | 37.51 ± 16.26 | 52.92 ± 20.88 | 37.25 ± 21.72 | 35.553 | <0.001*** |
| Number of functions endorsed | 14.51 ± 4.88 | 19.47 ± 5.75 | 15.13 ± 7.49 | 23.93 | <0.001*** |
| Number of body parts injured | 3.72 ± 2.42 | 3.86 ± 2.29 | 3.38 ± 1.76 | 0.746 | 0.475 |
| **Last month** | | | | | |
| Total functional scores | 37.12 ± 18.25 | 51.98 ± 25.99 | 40.78 ± 21.83 | 15.449 | <0.001*** |
| Number of functions endorsed | 13.93 ± 5.71 | 18.66 ± 6.92 | 15.54 ± 7.26 | 12.585 | <0.001*** |
| Number of body parts injured last month | 2.83 ± 1.96 | 2.42 ± 1.47 | 2.61 ± 1.78 | 1.224 | 0.296 |

*$p$ < 0.05; **$p$ <0.01; ***$p$ <0.001.

[a] The $\chi^2$ was obtained by a chi-square test.

**Table S3: Demographic data and NSSI characteristics between three diagnoses**

| | MDD (N = 167) | BD (N = 89) | ED (N = 48) | *F/χ2* | *p* |
|---|---|---|---|---|---|
| **Sex (F/M)** | 37/130 | 22/67 | 2/46 | 9.221[a] | 0.010* |
| **Age** | 18.35±2.26 | 19.58±2.59 | 18.31±2.05 | 8.932 | 0.001** |
| **SAS** | 59.54±14.26 | 62.51±15.41 | 58.75±14.31 | 1.514 | 0.222 |
| **SDS** | 70.73±14.09 | 71.26±12.95 | 58.75±14.31 | 0.567 | 0.568 |
| **ASLEC** | 48.61±21.93 | 51.46±24.36 | 57.27±20.65 | 2.815 | 0.061 |
| **FAD** | 148.23±22.30 | 147.58±22.11 | 146.54±22.54 | 0.894 | 0.894 |
| **CTQ** | 47.43±14.15 | 50.10±16.17 | 47.65±14.06 | 0.988 | 0.374 |
| **SSRS** | 29.16±6.65 | 30.65±6.34 | 29.13±6.60 | 1.638 | 0.196 |
| **CIAS** | 43.87±9.54 | 42.21±13.49 | 39.54±11.56 | 2.929 | 0.055 |
| **NEOFFI** | | | | | |
| Neuroticism | 37.63±5.85 | 37.28±6.67 | 36.35±6.70 | 0.456 | 0.456 |
| Extraversion | 21.85±6.49 | 22.23±7.28 | 22.02±6.73 | 0.886 | 0.886 |
| Openness to Experience | 34.15±6.45 | 34.02±5.84 | 32.15±6.71 | 0.145 | 0.145 |
| Agreeableness | 32.51±5.35 | 32.46±5.00 | 32.63±5.31 | 0.984 | 0.984 |
| Conscientiousness | 29.24±6.84 | 28.31±7.53 | 30.88±7.80 | 1.982 | 0.140 |
| NSSI Frequency (Repeated/ Occasional) | 101/66 | 56/33 | 21/27 | 5.289[a] | 0.071 |
| Self-related factor scores | 28.07 ± 12.79 | 30.65 ± 13.26 | 25.81 ± 10.62 | 3.704 | 0.025* |
| Self-related factor loading | 0.046 ± 0.032 | 0.043 ± 0.033 | 0.027 ± 0.028 | 6.822 | <0.001*** |
| Social-related factor scores | 12.46 ± 9.08 | 14.57 ± 11.22 | 17.23 ± 10.95 | 5.280 | 0.005** |
| Social-related factor loading | 0.029 ± 0.033 | 0.038 ± 0.043 | 0.053 ± 0.041 | 7.800 | <0.001*** |
| Suicidal Ideation (Yes/No) | 139/28 | 74/15 | 31/17 | 8.847[a] | 0.012* |
| Suicidal Attempt (Yes/No) | 94/73 | 62/26 | 22/26 | 8.697[a] | 0.013* |
| Age of onset | 15.28 ± 2.83 | 15.19 ± 3.87 | 14.74 ± 3.15 | 0.509 | 0.602 |
| **At onset** | | | | | |
| Total functional scores | 40.57 ± 18.96 | 45.40 ± 22.19 | 43.38 ± 19.74 | 0.831 | 0.277 |
| Number of functions endorsed | 15.40 ± 6.02 | 17.11 ± 6.11 | 17.40 ± 6.02 | 3.884 | 0.034* |
| Number of body parts injured | 3.80 ± 2.42 | 3.40 ± 1.96 | 3.94 ± 2.31 | 0.825 | 0.314 |

M/F, Male/Female; MDD, major depressive disorder; BD, bipolar disorder; ED, eating disorders; SAS, Self-Rating Anxiety Scale; SDS, Self-Rating Depression Scale; ASLEC, Adolescent Self-Rating Life Events Checklist; FAD, Family Assessment Device; CTQ, Childhood Trauma Questionnaire; SSRS, Social Support Rating Scale; CIAS, Chinese Internet Addiction Scale; NEOFFI, NEO Five-Factor Inventory. *$p$ <0.05; **$p$ <0.01; ***$p$ <0.001.

[a] The $\chi^2$ was obtained by a chi-square test.

#### Table S4: Functional connectivity related to factors of NSSI functions

| Factors | Region name | Region name | *r* | *p* |
|---|---|---|---|---|
| **Self-related** | Amygdala (R) | Dorsolateral area 6 (R) | -0.29 | 1.73E-04 |
| | Amygdala (R) | Area 4 (trunk region) (R) | -0.30 | 1.20E-04 |
| | Amygdala (R) | TE1.0 and TE1.2 (L) | -0.31 | 5.24E-05 |
| | Amygdala (R) | Caudal area 35/36 (L) | -0.29 | 1.84E-04 |
| | Amygdala (R) | Area 1/2/3 (tongue and larynx region) (R) | -0.28 | 3.28E-04 |
| | Amygdala (R) | Area1/2/3 (trunk region) (L) | -0.28 | 2.81E-04 |
| | Amygdala (R) | Area1/2/3 (trunk region) (R) | -0.27 | 4.81E-04 |
| | Amygdala (R) | Hypergranular insula (L) | -0.27 | 4.36E-04 |
| | Amygdala (R) | Hypergranular insula (R) | -0.26 | 7.33E-04 |
| | Amygdala (R) | Caudal lingual gyrus (R) | -0.27 | 5.27E-04 |
| | Amygdala (R) | Caudal cuneus gyrus (L) | -0.28 | 2.77E-04 |
| | Amygdala (R) | Ventromedial parietooccipital sulcus (L) | -0.26 | 9.89E-04 |
| | Amygdala (R) | Ventromedial parietooccipital sulcus (R) | -0.26 | 9.29E-04 |
| | Amygdala (R) | Occipital polar cortex (L) | -0.26 | 8.04E-04 |
| | Amygdala (R) | Inferior occipital gyrus (L) | -0.26 | 9.01E-04 |
| | Amygdala (R) | Medial superior occipital gyrus (L) | -0.26 | 6.67E-04 |
| | Amygdala (L) | Putamen (R) | -0.26 | 9.59E-04 |
| | Caudal cuneus gyrus (L) | Caudoposterior superior temporal sulcus (R) | -0.27 | 5.51E-04 |
| | Caudal cuneus gyrus (L) | Opercular area 44 (R) | -0.260 | 8.437e-04 |
| | Putamen (L) | Ventromedial parietooccipital sulcus (L) | -0.26 | 9.92E-04 |
| | Putamen (L) | Ventromedial parietooccipital sulcus (R) | -0.26 | 6.58E-04 |
| | Putamen (R) | Caudal cuneus gyrus (L) | -0.28 | 3.59E-04 |
| | Putamen (R) | Ventromedial parietooccipital sulcus (L) | -0.26 | 7.96E-04 |
| **Social-related** | Caudolateral area 20 (R) | Hippocampus (L) | -0.26 | 7.76E-04 |
| | Dorsal area 9/46 (L) | Lateral area 5 (R) | -0.27 | 3.89E-04 |
| | Dorsal area 9/46 (L) | Postcentral area 7 (L) | -0.27 | 4.42E-04 |
| | Dorsolateral area 6 (R) | Hypergranular insula (R) | -0.28 | 2.35E-04 |
| | Dorsolateral area 8 (L) | Lateroventral area37 (L) | -0.28 | 2.92E-04 |
| | Ventrolateral area 8 (L) | Lateroventral area37 (L) | -0.26 | 7.72E-04 |

L, left; R, right.

**Table S5: Cross-diagnostic comparison of functional connectivity related to factors**

| Factors | Region name | Region name | $F_{(1,160)}$ | $p$ FDR cor |
|---|---|---|---|---|
| **Self-related** | Amygdala (R) | Dorsolateral area 6 (R) | 0.211 | 0.823 |
| | Amygdala (R) | Area 4 (trunk region) (R) | 0.183 | 0.823 |
| | Amygdala (R) | TE1.0 and TE1.2 (L) | 0.236 | 0.689 |
| | Amygdala (R) | Caudal area 35/36 (L) | 0.189 | 0.706 |
| | Amygdala (R) | Area 1/2/3 (tongue and larynx region) (R) | 0.204 | 0.793 |
| | Amygdala (R) | Area1/2/3 (trunk region) (L) | 0.201 | 0.802 |
| | Amygdala (R) | Area1/2/3 (trunk region) (R) | 0.194 | 0.823 |
| | Amygdala (R) | Hypergranular insula (L) | 0.232 | 0.191 |
| | Amygdala (R) | Hypergranular insula (R) | 0.218 | 0.31 |
| | Amygdala (R) | Caudal lingual gyrus (R) | 0.193 | 0.823 |
| | Amygdala (R) | Caudal cuneus gyrus (L) | 0.196 | 0.83 |
| | Amygdala (R) | Ventromedial parietooccipital sulcus (L) | 0.197 | 0.823 |
| | Amygdala (R) | Ventromedial parietooccipital sulcus (R) | 0.198 | 0.823 |
| | Amygdala (R) | Occipital polar cortex (L) | 0.180 | 0.961 |
| | Amygdala (R) | Inferior occipital gyrus (L) | 0.190 | 0.823 |
| | Amygdala (R) | Medial superior occipital gyrus (L) | 0.184 | 0.961 |
| | Amygdala (L) | Putamen (R) | 0.224 | 0.706 |
| | Caudal cuneus gyrus (L) | Caudoposterior superior temporal sulcus (R) | 0.204 | 0.31 |
| | Caudal cuneus gyrus (L) | Opercular area 44 (R) | 0.196 | 0.706 |
| | Putamen (L) | Ventromedial parietooccipital sulcus (L) | 0.177 | 0.706 |
| | Putamen (L) | Ventromedial parietooccipital sulcus (R) | 0.180 | 0.823 |
| | Putamen (R) | Caudal cuneus gyrus (L) | 0.186 | 0.823 |
| | Putamen (R) | Ventromedial parietooccipital sulcus (L) | 0.184 | 0.706 |
| **Social-related** | Caudolateral area 20 (R) | Hippocampus (L) | 0.207 | 0.992 |
| | Dorsal area 9/46 (L) | Lateral area 5 (R) | 0.245 | 0.635 |
| | Dorsal area 9/46 (L) | Postcentral area 7 (L) | 0.239 | 0.635 |
| | Dorsolateral area 6 (R) | Hypergranular insula (R) | 0.245 | 0.635 |
| | Dorsolateral area 8 (L) | Lateroventral area37 (L) | 0.223 | 0.992 |
| | Ventrolateral area 8 (L) | Lateroventral area37 (L) | 0.247 | 0.635 |

L, left; R, right.

### III. Supplementary Figures

#### Figure S1: Stability and generalizability metrics of the two-factor model

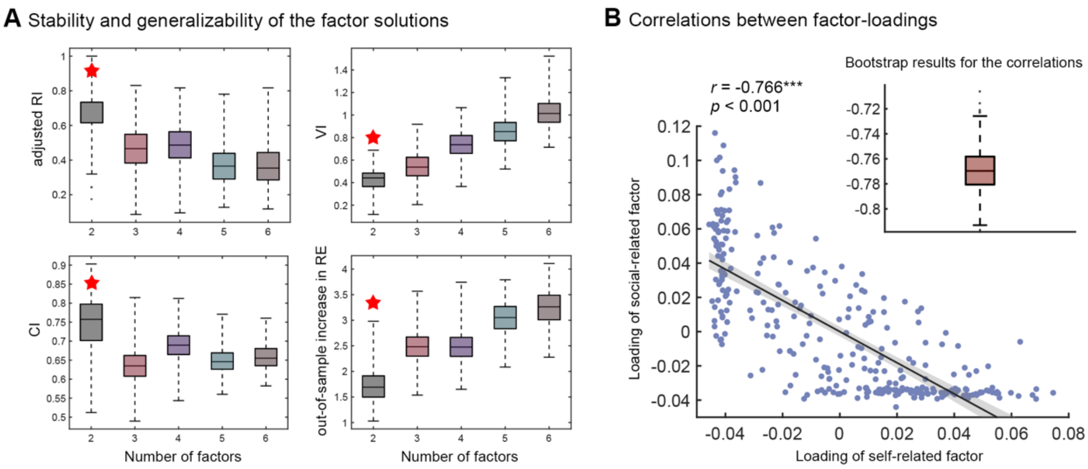

**(A)** The three indices used to examine the stability and generalizability of the factor model. Higher values for adjusted RI and CI (upper row) denote greater stability. Conversely, lower values for VI and an increase in out-of-sample RE (bottom row) indicate improved stability and generalizability, respectively. **(B)** The scatter plot shows the correlation between the two factor loadings, with the correlation coefficient tested by bootstrap. For each box plot, the box illustrates the Standard Error of the Mean (SEM, centered on the mean), whiskers denote the 5% and 95% values, and the horizontal line signifies the median. The star represents the optimal number of factors. adjusted RI, adjusted Rand index; VI, variation of information; CI, concordance index; RE, reconstruction error. ***$p < 0.001$.

**Figure S2: Determination of optimal cluster number and membership cutoff of Fuzzy C-means clustering**

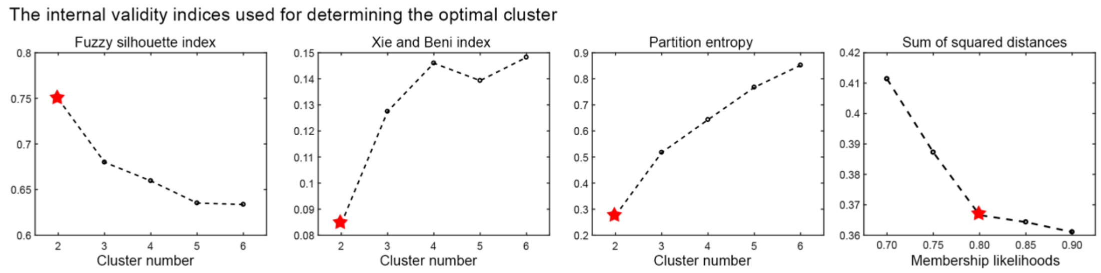

The fuzzy silhouette index (SI), Xie and Beni index (XB), and partition entropy (PE) were calculated at range of 2 to 6 as internal validity indices to determine the optimal number of clusters. Higher SI and lower XB and PE indicated better clustering quality. To choose the cutoff to define the optimal membership discrimination, the elbow method was used at range of 70% to 90% by identifying the elbow point, where the rate of decrease in explained variance slows significantly. The elbow point represented a balance between maximizing cluster compactness and minimizing the number of clusters. The star represents the optimal number of factors.

#### Figure S3: Self–Social dimensional model of NSSI functions using revised OSI-F

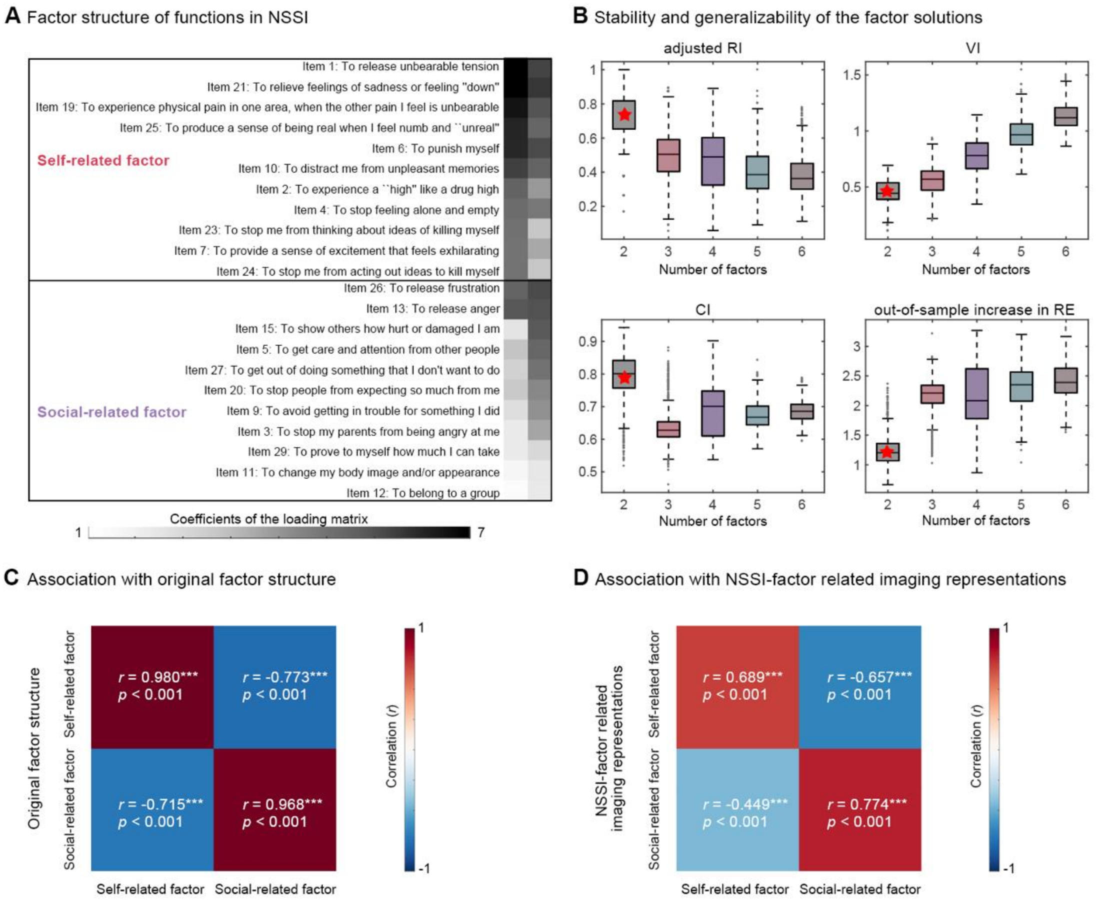

**(A)**The two-factor structure of revised OSI-F (22 items) output by OPNMF. The self-related factor and a social-related factor contain 11 and 11 items respectively. **(B)** The three indices used to examine the stability and generalizability of the factor model. Higher values for adjusted RI and CI (upper row) denote greater stability. Conversely, lower values for VI and an increase in out-of-sample RE (bottom row) indicate improved stability and generalizability, respectively. For each box plot, the box illustrates the Standard Error of the Mean (SEM, centered on the mean), whiskers denote the 5% and 95% values, and the horizontal line signifies the median. The star represents the optimal number of factors. **(C)** The relationship between the factor structures derived from the revised OSI-F (22 items) and the original OSI-F (29 items). **(D)** The relationship between the two factors and the previously identified rsFC characteristics. Red indicating positive correlations and blue indicating negative correlations. adjusted RI, adjusted Rand index; VI, variation of information; CI, concordance index; RE, reconstruction error. ***$p < 0.001$.

#### Figure S4: Self–Social dimensional model in patients with repeated NSSI

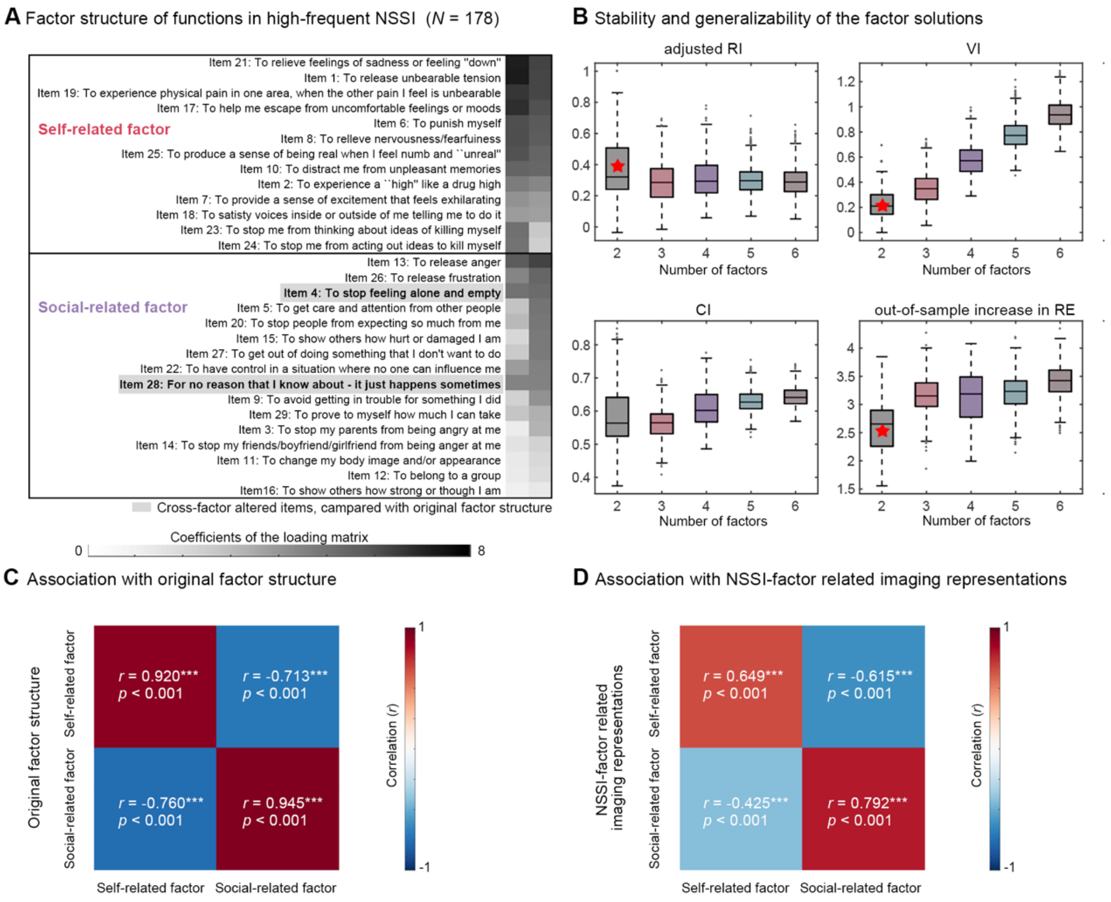

**(A)** The two-factor model of NSSI functions in patients with repeated-NSSI (NSSI behaviors occurred for at least 5 days in the past year). Items that differ from the original factor structure are bolded and shaded. **(B)** The three indices used to examine the stability and generalizability of the factor model. Higher values for adjusted RI and CI (upper row) denote greater stability. Conversely, lower values for VI and an increase in out-of-sample RE (bottom row) indicate improved stability and generalizability, respectively. For each box plot, the box illustrates the Standard Error of the Mean (SEM, centered on the mean), whiskers denote the 5% and 95% values, and the horizontal line signifies the median. The star represents the optimal number of factors. **(C)** The relationship between the factor structures derived from the repeated-NSSI patients (N = 178) and full samples (N = 204). **(D)** The relationship between the two factors and the previously identified rsFC characteristics. Red indicating positive correlations and blue indicating negative correlations. adjusted RI, adjusted Rand index; VI, variation of information; CI, concordance index; RE, reconstruction error. ****p* < 0.001.

#### Figure S5: Self–Social dimensional model in patients with occasional NSSI

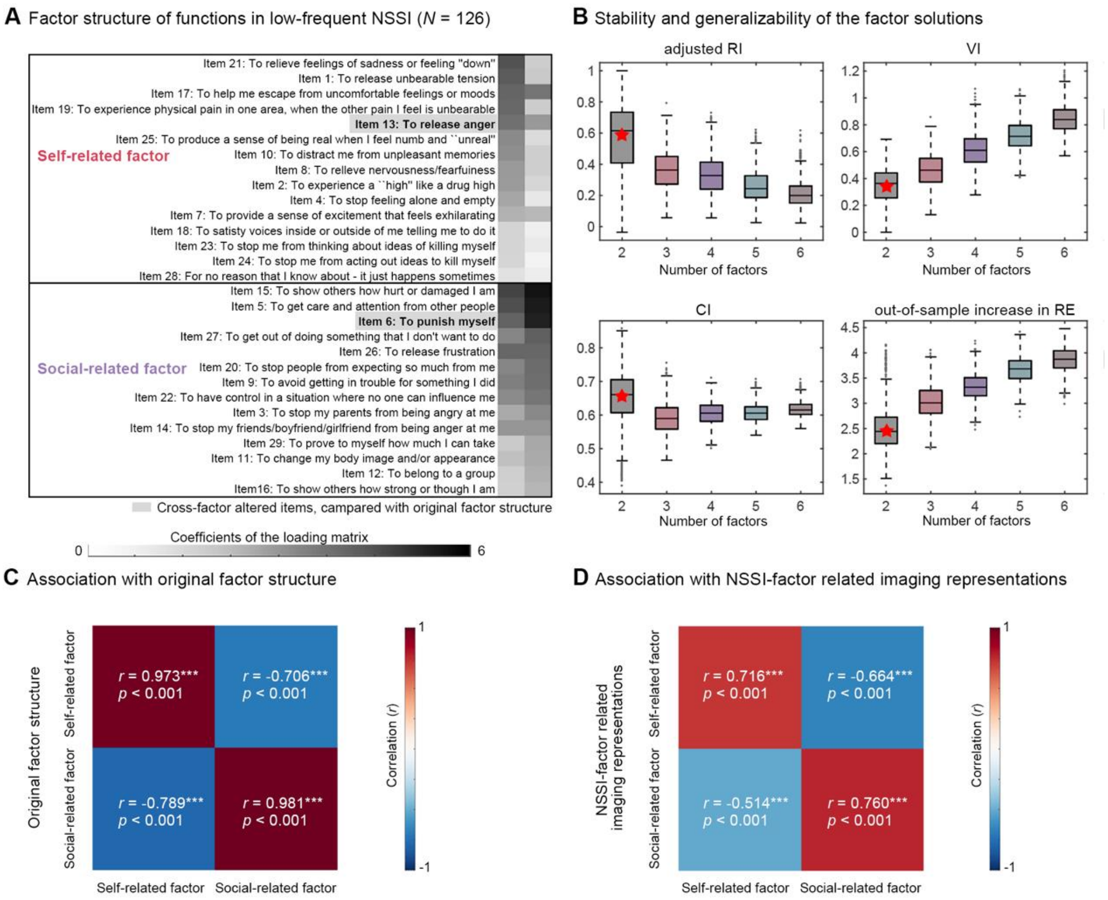

**(A)** The two-factor model of NSSI functions in patients with occasional-NSSI (NSSI behaviors occurred on at least one day but fewer than 5 days). Items that differ from the original factor structure are bolded and shaded. **(B)** The three indices used to examine the stability and generalizability of the factor model. Higher values for adjusted RI and CI (upper row) denote greater stability. Conversely, lower values for VI and an increase in out-of-sample RE (bottom row) indicate improved stability and generalizability, respectively. For each box plot, the box illustrates the Standard Error of the Mean (SEM, centered on the mean), whiskers denote the 5% and 95% values, and the horizontal line signifies the median. The star represents the optimal number of factors. **(C)** The relationship between the factor structures derived from the repeated-NSSI patients (N = 126) and full samples (N = 204). **(D)** The relationship between the two factors and the previously identified rsFC characteristics. Red indicating positive correlations and blue indicating negative correlations. adjusted RI, adjusted Rand index; VI, variation of information; CI, concordance index; RE, reconstruction error. ***$p < 0.001$.

#### Figure S6: Functional connectivity related to two factors

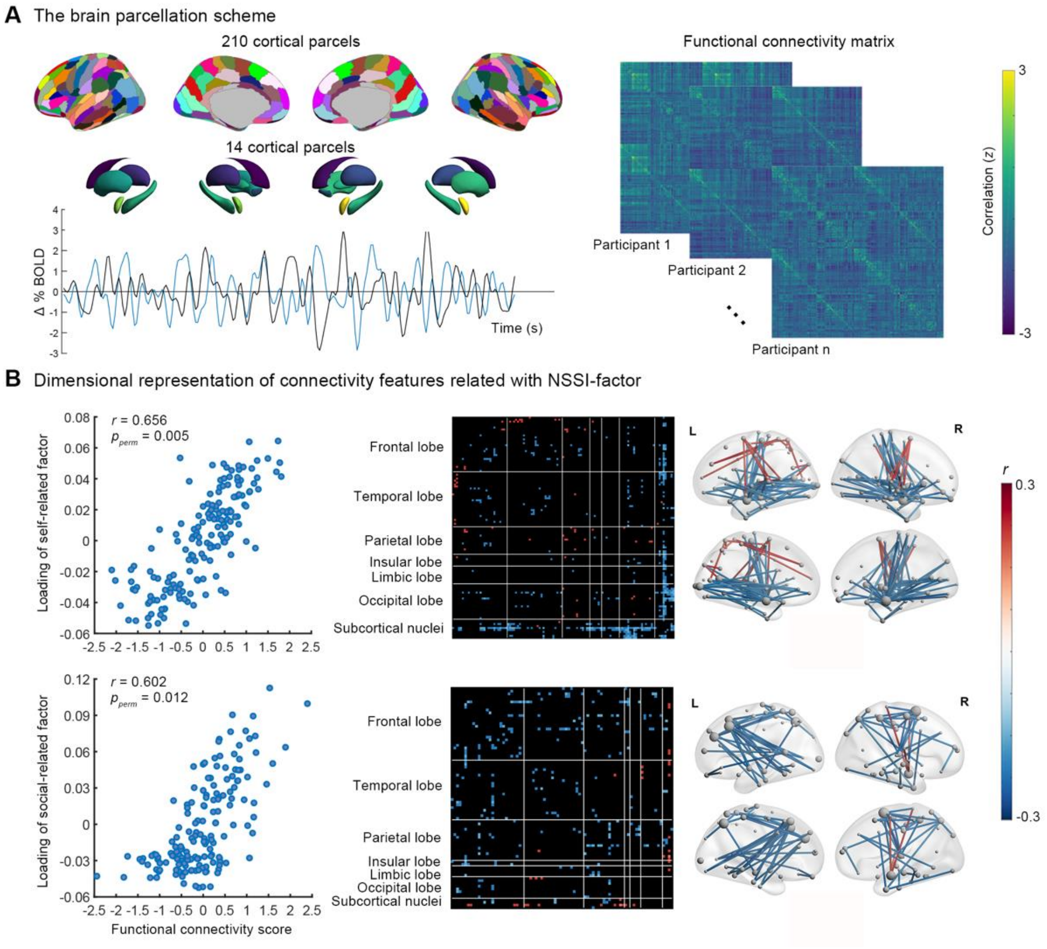

**(A)** Using a brain parcellation template containing 210 cortical areas and 14 subcortical areas, time series were extracted to construct a functional connectivity matrix. **(B)** Scatterplots (left) illustrate the association between connectivity scores and the loading of functional factor scores for each RCC dimension across participants. The heat map is labeled by 224 brain regions (y-axis, grouped into six cortical and one subcortical region) and factor scores (x-axis), showing the mean correlation between rsFC features and two scores. Red indicates positive correlations, while blue indicates negative correlations ($p < 0.05$, FDR corrected). The glass brain illustrates the neuroanatomical distribution of rsFC features in the chord plot. Each node represents a brain region, with larger nodes indicating more connections. The chord plot (right) highlights the most significant rsFC features ($p < 0.001$, FDR corrected).

#### Figure S7: The workflow of regularized canonical correlation analysis (RCC)

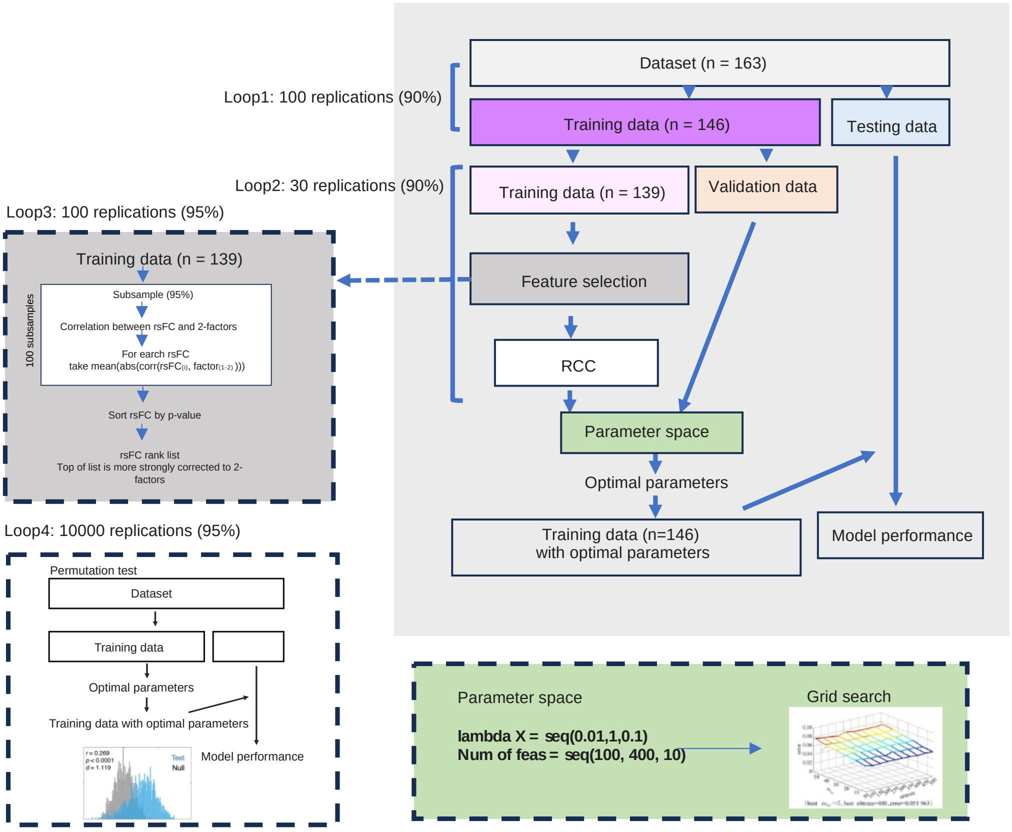

The light gray area represents the main RCC process. The dark gray area, bright blue area, and green area represent the details correspond to the feature selection, model performance, and parameter space in the main process, respectively.

#### Reference

1 Chen J, Patil KR, Weis S, Sim K, Nickl-Jockschat T, Zhou J *et al.* Neurobiological Divergence of the Positive and Negative Schizophrenia Subtypes Identified on a New Factor Structure of Psychopathology Using Non-negative Factorization: An International Machine Learning Study. *Biological Psychiatry* 2020; **87**: 282–293.

2 Hubert L, Arabie P. Comparing partitions. *Journal of classification* 1985; **2**: 193–218.

3 Meilă M. Comparing clusterings—an information based distance. *Journal of multivariate analysis* 2007; **98**: 873–895.

4 Raguideau S, Plancade S, Pons N, Leclerc M, Laroche B. Inferring aggregated functional traits from metagenomic data using constrained non-negative matrix factorization: Application to fiber degradation in the human gut microbiota. *PLoS computational biology* 2016; **12**: e1005252.

5 Campello RJ, Hruschka ER. A fuzzy extension of the silhouette width criterion for cluster analysis. *Fuzzy Sets and Systems* 2006; **157**: 2858–2875.

6 Xie XL, Beni G. A validity measure for fuzzy clustering. *IEEE Transactions on Pattern Analysis & Machine Intelligence* 1991; **13**: 841–847.

7 Bezdek JC, Bezdek JC. Objective function clustering. *Pattern recognition with fuzzy objective function algorithms* 1981; : 43–93.

8 Shi C, Wei B, Wei S, Wang W, Liu H, Liu J. A quantitative discriminant method of elbow point for the optimal number of clusters in clustering algorithm. J Wireless Com Network 31: 1–16. 2021.

9 Yan C-G, Wang X-D, Zuo X-N, Zang Y-F. DPABI: data processing & analysis for (resting-state) brain imaging. *Neuroinformatics* 2016; **14**: 339–351.

10 Friston KJ, Williams S, Howard R, Frackowiak RS, Turner R. Movement-related effects in fMRI time-series. *Magnetic resonance in medicine* 1996; **35**: 346–355.

11 Buch AM, Vértes PE, Seidlitz J, Kim SH, Grosenick L, Liston C. Molecular and network-level mechanisms explaining individual differences in autism spectrum disorder. *Nat Neurosci* 2023; **26**: 650–663.

12 Klonsky ED, Glenn CR. Assessing the Functions of Non-suicidal Self-injury: Psychometric Properties of the Inventory of Statements About Self-injury (ISAS). *J Psychopathol Behav Assess* 2009; **31**: 215–219.

13 Rodav O, Levy S, Hamdan S. Clinical characteristics and functions of non-suicide self-injury in youth. *Eur psychiatr* 2014; **29**: 503–508.

14 Case JAC, Burke TA, Siegel DM, Piccirillo ML, Alloy LB, Olino TM. Functions of Non-Suicidal Self-Injury in Late Adolescence: A Latent Class Analysis. *Archives of Suicide Research* 2020; **24**: S165–S186.
